# Genetic and Social Determinants of Renin-Angiotensin-Aldosterone System Inhibitor-Induced Angioedema: A Precision Medicine Health Equity Study

**DOI:** 10.64898/2026.01.06.26343530

**Authors:** Nanase Toda, Tanushree Haldar, Craig C. Teerlink, Donglei Hu, Peter Danilov, Scott Huntsman, Meng Lu, Philip S. Tsao, Catherine Tcheandjieu, Carlos Iribarren, Adam Bress, Julie A. Lynch, Elad Ziv, Akinyemi Oni-Orisan

## Abstract

Angioedema is a life-threatening adverse drug reaction associated with renin-angiotensin-aldosterone system (RAAS) inhibitors, characterized by localized swelling in the deep layers of the skin. Well-established evidence indicates an up to fivefold higher incidence of RAAS inhibitor-induced angioedema in self-identified Black patients compared to White patients. The mechanisms underlying this health disparity remain poorly understood and are often attributed to race, a poor proxy for interindividual genetic similarity and social stressors. Here, we investigate the genetic and social determinants of RAAS inhibitor-induced angioedema as well as the etiology of this racial difference. In particular, we (1) discovered *OTULINL* and *CRABP1* as novel loci for RAAS inhibitor-induced angioedema, (2) confirmed the importance of bradykinin for this adverse drug reaction, (3) reported the first significant genome-wide association in self-identified Black participants, (4) identified alcohol use as an important social determinant, (5) demonstrated the strong role of variants enriched in 1000 Genomes African superpopulation-like genomes as the driver of racially differential angioedema risk, and (6) demonstrated the combined role of polygenic effect size and allele frequency differences in explaining these racial differences. Our results suggest that a clinical precision medicine tool may more precisely predict for whom RAAS inhibitors should be avoided (to prevent angioedema) compared to using race. These findings ultimately underscore the value of an evidence-based approach to removing race from treatment guidelines, which carries less potential harm than other removal strategies.

## Introduction

Renin-angiotensin-aldosterone system (RAAS) inhibitors are life-saving medications^1^ prescribed to over 30 million adults in the United States for hypertension alone^2^, a number that underestimates total exposure given their additional indications in heart failure, chronic kidney disease, and other conditions^1^. Although generally well tolerated^3^, 0.1–0.7% of users develop angioedema—a life-threatening condition characterized by localized swelling in the deep layers of the skin^4^. Given the widespread use of these medications, this prevalence translates to hundreds of thousands of affected individuals, underscoring the substantial public health impact of this adverse drug reaction (ADR). Notably, the incidence of RAAS inhibitor-induced angioedema is consistently up to fivefold higher in self-identified Black patients compared to those identifying as White^5–7^. This is considered a health disparity per the most recent National Institute on Minority Health and Health Disparities (NIMHD) definition: “a health difference, on the basis of one or more health outcomes, that adversely affects disadvantaged populations^8^.”

Despite acknowledgement of this well-established differential drug response in prior clinical guidelines for heart failure management^9–11^, the most recent guidelines have omitted this important disparity^12^. This exclusion was not accompanied with any new evidence justifying its omisison. Although race is a poor marker for interindividual genetic similarity and a crude surrogate of social stressors^13^, simply ignoring its role in RAAS inhibitor-induced angioedema may perpetuate this health disparity^14^. Additionally, prior guidelines did not provide health care providers with actionable strategies to address this disparity. Elucidating the complex genetic and social determinants underlying this differential drug response observation offers an evidence-based, alternative approach that has the potential to address this disparity while simultaneously obviating the use of race as a proxy in medicine.

Although there have been some genome-wide association studies (GWASs) for RAAS inhibitor-induced angioedema^15–18^, the elucidation of determinants underlying differential risk between Black and White patients has not been explicitly explored. A major barrier to addressing this knowledge gap has been the Eurocentric bias of study populations in pharmacogenetic research^19,20^. However, the recent emergence of large, racially diverse datasets that adequately capture both genetic data and social determinants of health is enabling enhanced research for groups traditionally understudied in pharmacogenetics^21,22^. These resources promise to improve drug safety outcomes for these groups. Therefore, we sought to identify genetic and social factors underlying the differential RAAS inhibitor-induced angioedema risk between Black and White patient groups, leveraging some of the country’s most racially diverse electronic health record (EHR)-linked biobanks.

## Results

### Characteristics of the Study Population

The full study population included 476 744 RAAS inhibitor users (comprising 6872 RAAS inhibitor-induced angioedema cases and 469 872 controls). This includes 92 282 total self-identified Black participants and 440 039 participants who self-identified as White. The matched study population comprised 4726 cases. See Extended Data Fig. 1 for more details on sample size, including numbers by cohort and at each stage of the study population generation.

In the matched study population, 9185 participants (18%) were female and 47 333 (91%) were angiotensin-converting enzyme (ACE) inhibitor users at index date. Among index pharmacy records, the most common drug type was lisinopril (84%). Table 1 provides more details on characteristics at index date for Black and White participants in each cohort. Extended Data Table 1 shows characteristics for the 19 516 participants with >5% 1000 Genomes African superpopulation (1KG-AFR)-like genetic similarity.

**Table 1.**
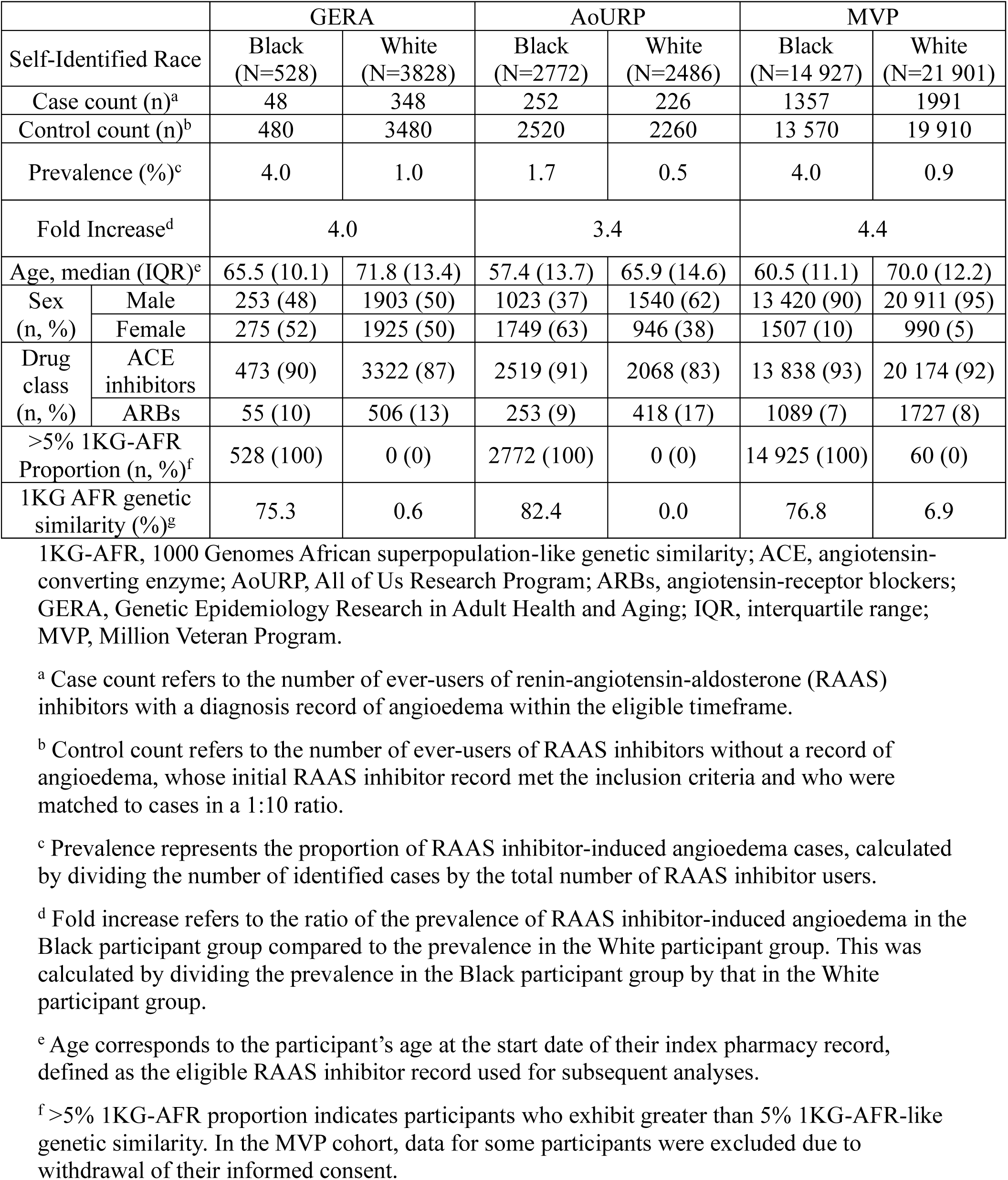
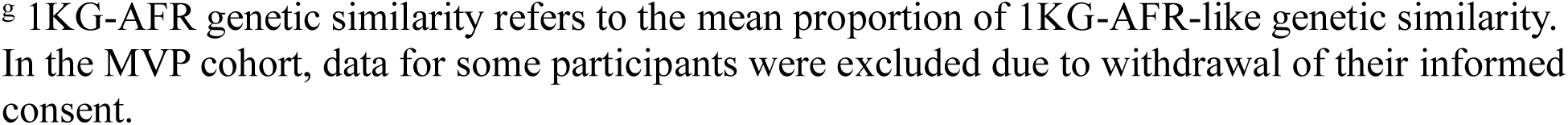
Demographic and clinical characteristics in the matched study population.

|  |  | GERA |  | AoURP |  | MVP |  |
| --- | --- | --- | --- | --- | --- | --- | --- |
| Self-Identified Race |  | Black<br>(N=528) | White<br>(N=3828) | Black<br>(N=2772) | White<br>(N=2486) | Black<br>(N=14 927) | White<br>(N=21 901) |
| Case count (n)<sup>a</sup> |  | 48 | 348 | 252 | 226 | 1357 | 1991 |
| Control count (n)<sup>b</sup> |  | 480 | 3480 | 2520 | 2260 | 13 570 | 19 910 |
| Prevalence (%)<sup>c</sup> |  | 4.0 | 1.0 | 1.7 | 0.5 | 4.0 | 0.9 |
| Fold Increase<sup>d</sup> |  | 4.0 |  | 3.4 |  | 4.4 |  |
| Age, median (IQR)<sup>e</sup> |  | 65.5 (10.1) | 71.8 (13.4) | 57.4 (13.7) | 65.9 (14.6) | 60.5 (11.1) | 70.0 (12.2) |
| Sex<br>(n, %) | Male | 253 (48) | 1903 (50) | 1023 (37) | 1540 (62) | 13 420 (90) | 20 911 (95) |
|  | Female | 275 (52) | 1925 (50) | 1749 (63) | 946 (38) | 1507 (10) | 990 (5) |
| Drug<br>class<br>(n, %) | ACE<br>inhibitors | 473 (90) | 3322 (87) | 2519 (91) | 2068 (83) | 13 838 (93) | 20 174 (92) |
|  | ARBs | 55 (10) | 506 (13) | 253 (9) | 418 (17) | 1089 (7) | 1727 (8) |
| >5% 1KG-AFR<br>Proportion (n, %)<sup>f</sup> |  | 528 (100) | 0 (0) | 2772 (100) | 0 (0) | 14 925 (100) | 60 (0) |
| 1KG AFR genetic<br>similarity (%)<sup>g</sup> |  | 75.3 | 0.6 | 82.4 | 0.0 | 76.8 | 6.9 |

1KG-AFR, 1000 Genomes African superpopulation-like genetic similarity; ACE, angiotensin-converting enzyme; AoURP, All of Us Research Program; ARBs, angiotensin-receptor blockers; GERA, Genetic Epidemiology Research in Adult Health and Aging; IQR, interquartile range; MVP, Million Veteran Program.
<sup>a</sup> Case count refers to the number of ever-users of renin-angiotensin-aldosterone (RAAS) inhibitors with a diagnosis record of angioedema within the eligible timeframe.
<sup>b</sup> Control count refers to the number of ever-users of RAAS inhibitors without a record of angioedema, whose initial RAAS inhibitor record met the inclusion criteria and who were matched to cases in a 1:10 ratio.
<sup>c</sup> Prevalence represents the proportion of RAAS inhibitor-induced angioedema cases, calculated by dividing the number of identified cases by the total number of RAAS inhibitor users.
<sup>d</sup> Fold increase refers to the ratio of the prevalence of RAAS inhibitor-induced angioedema in the Black participant group compared to the prevalence in the White participant group. This was calculated by dividing the prevalence in the Black participant group by that in the White participant group.
<sup>e</sup> Age corresponds to the participant's age at the start date of their index pharmacy record, defined as the eligible RAAS inhibitor record used for subsequent analyses.
<sup>f</sup> >5% 1KG-AFR proportion indicates participants who exhibit greater than 5% 1KG-AFR-like genetic similarity. In the MVP cohort, data for some participants were excluded due to withdrawal of their informed consent.
<sup>g</sup> 1KG-AFR genetic similarity refers to the mean proportion of 1KG-AFR-like genetic similarity. In the MVP cohort, data for some participants were excluded due to withdrawal of their informed consent.

### Characteristics of RAAS Inhibitor-Induced Angioedema

The prevalence of RAAS inhibitor-induced angioedema among self-identified Black participants was 4.0%, 1.7%, and 4.0% in the Genetic Epidemiology Research in Adult Health and Aging (GERA), All of Us Research Program (AoURP), and Million Veteran Program (MVP) cohorts, respectively. In comparison, the prevalence among self-identified White participants was 1.0%, 0.5%, and 0.9% in the respective cohorts. The corresponding fold increases in prevalence among Black participants compared to White Participants were 4.0, 3.4, and 4.4.

Among established clinical and demographic factors, Black race had the strongest independent association with RAAS inhibitor-induced angioedema (odds ratio [OR] = 5.16, 95% CI 3.92-6.78, *P* = 1.6 × 10^-30^ in GERA; OR = 4.01, CI 3.43-4.70, *P* = 3.2 × 10^-67^ in AoURP; OR = 5.58, CI 5.25-5.95, *P* = 1.2 × 10^-621^ in MVP, Table 2). Moreover, ACE inhibitor use had a strong effect relative to angiotensin-receptor blocker (ARB) use in the MVP and AoURP cohorts (OR = 3.22, CI 2.60-4.00, P = 8.6 × 10^-28^ in AoURP; OR = 3.32, CI 2.89-3.81, *P* = 1.3 × 10^-65^ in MVP) but not in GERA (OR = 0.84, CI 0.65-1.08, *P* = 0.17). Age was also a strong predictor of the outcome (OR = 1.04 per year, CI 1.03-1.05, *P* = 1.1 × 10^-22^ in GERA; OR = 1.03, CI 1.02-1.04, *P* = 6.4 × 10^-19^ in AoURP; OR = 1.05, CI 1.05-1.05, *P* = 8.7 × 10^-219^ in MVP).

**Table 2.**
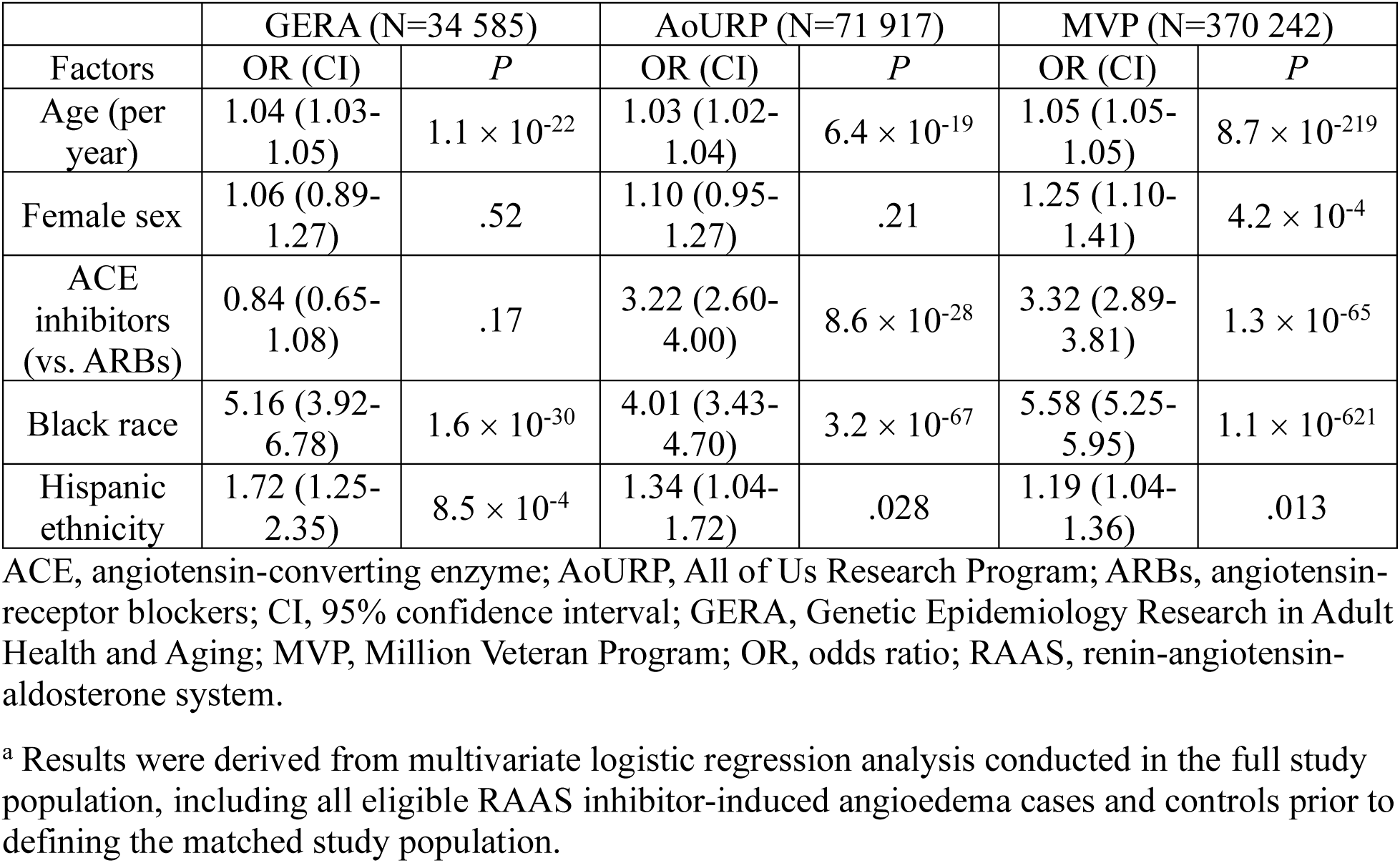
Clinical and demographic predictors of RAAS inhibitor-induced angioedema^a^.

| Factors | GERA (N=34 585) |  | AoURP (N=71 917) |  | MVP (N=370 242) |  |
| --- | --- | --- | --- | --- | --- | --- |
|  | OR (CI) | *P* | OR (CI) | *P* | OR (CI) | *P* |
| Age (per year) | 1.04 (1.03-1.05) | $1.1 \times 10^{-22}$ | 1.03 (1.02-1.04) | $6.4 \times 10^{-19}$ | 1.05 (1.05-1.05) | $8.7 \times 10^{-219}$ |
| Female sex | 1.06 (0.89-1.27) | .52 | 1.10 (0.95-1.27) | .21 | 1.25 (1.10-1.41) | $4.2 \times 10^{-4}$ |
| ACE inhibitors (vs. ARBs) | 0.84 (0.65-1.08) | .17 | 3.22 (2.60-4.00) | $8.6 \times 10^{-28}$ | 3.32 (2.89-3.81) | $1.3 \times 10^{-65}$ |
| Black race | 5.16 (3.92-6.78) | $1.6 \times 10^{-30}$ | 4.01 (3.43-4.70) | $3.2 \times 10^{-67}$ | 5.58 (5.25-5.95) | $1.1 \times 10^{-621}$ |
| Hispanic ethnicity | 1.72 (1.25-2.35) | $8.5 \times 10^{-4}$ | 1.34 (1.04-1.72) | .028 | 1.19 (1.04-1.36) | .013 |

ACE, angiotensin-converting enzyme; AoURP, All of Us Research Program; ARBs, angiotensin-receptor blockers; CI, 95% confidence interval; GERA, Genetic Epidemiology Research in Adult Health and Aging; MVP, Million Veteran Program; OR, odds ratio; RAAS, renin-angiotensin-aldosterone system.
<sup>a</sup> Results were derived from multivariate logistic regression analysis conducted in the full study population, including all eligible RAAS inhibitor-induced angioedema cases and controls prior to defining the matched study population.

### Social Determinants of RAAS Inhibitor-Induced Angioedema

Among available social determinants of health (SDOH) data, 18 factors in the GERA cohort and 29 factors in the AoURP cohort met the inclusion criteria for data analysis.

Of these included SDOH, five were significantly associated with the phenotype in the GERA cohort (Extended Data Table 2) and four in AoURP (Extended Data Table 3). Specifically, in the GERA cohort, marital status (OR = 2.80, CI 1.16-6.77, *P* = .023) was significantly associated with the phenotype in the self-identified Black participant group. In the self-identified White participant group, alcohol consumption (OR = 1.28 per alcoholic drink consumed daily, CI 1.13-1.46, *P* < .001), education level (OR = 0.87, CI 0.77-0.98, *P* = .023), income (OR = 1.20, CI 1.03-1.40, *P* = .022), and walking (OR = 0.96, CI 0.92-1.00, *P* = .037) were significantly associated with the phenotype. In the AoURP cohort, general quality of life (OR = 1.48, CI 1.06-2.06, *P* = .024), general satisfaction with social activities and relationships (OR = 0.70, CI 0.53-0.92, *P* = .011), general social role performance (OR = 1.43, CI 1.07-1.92, *P* = .018), and years lived at the current address (OR = 1.03, CI 1.01-1.05, *P* = .013) were significantly associated with the phenotype in the self-identified White participant group.

### Genetic Similarity and SDOH as Factors Underlying Racially Different RAAS Inhibitor-Induced Angioedema Prevalence

The association of Black race with RAAS inhibitor-induced angioedema risk (OR 5.16, *P* = 1.6 × 10^-30^ in GERA; OR = 4.01, *P* = 3.2 × 10^-67^ in AoURP) did not change substantively when SDOH was added to the model (OR 5.64, *P* = 4.5 × 10^-24^ in GERA; OR 3.94, *P* = 4.5 × 10^-57^ in AoURP; Extended Data Fig. 2 and Fig. 1). In contrast, the association was entirely abrogated when 1KG-AFR-like genetic similarity was added (OR 1.10, *P* = .81 in GERA; OR 0.89, *P* = .48 in AoURP; Extended Data Fig. 2 and Fig. 1).

**Figure 1.**
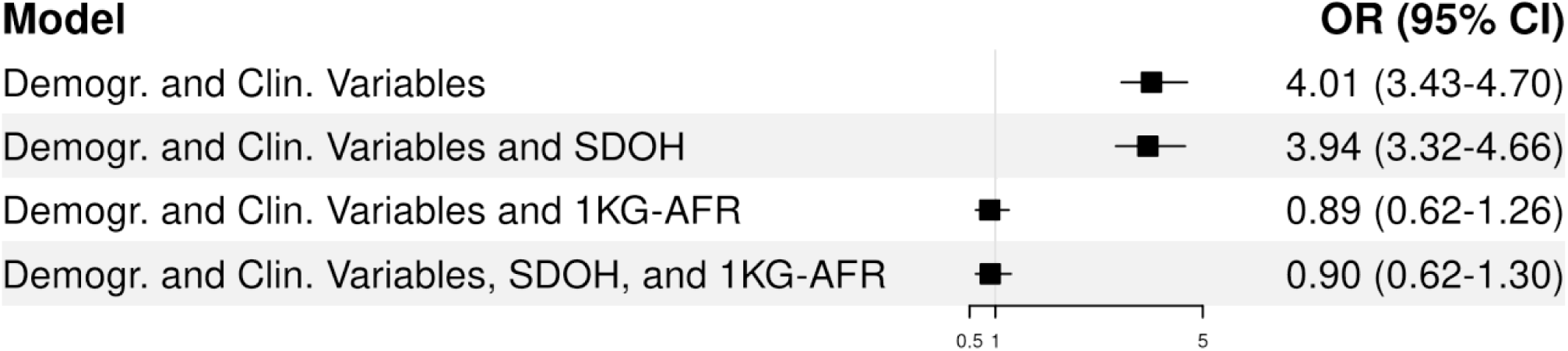
Association of Black race with RAAS inhibitor-induced angioedema risk in the AoURP cohort across multivariable logistic regression models 1KG-AFR, 1000 Genomes African superpopulation; AoURP, All of Us Research Program; clin., clinical; demogr., demographic; RAAS, renin-angiotensin-aldosterone system; SDOH, social determinants of health. The inclusion of SDOH minimally explained the observed association with Black race, while the addition of 1KG-AFR-like genetic similarity completely abrogated the race effect.

### Genome-Wide Variants of RAAS Inhibitor-Induced Angioedema

Association results from within-racial group meta-GWAS analyses are shown in Fig. 2 and Fig. 3. Genome-wide significance was observed for 50 SNVs (formerly SNP) from one locus in the self-identified Black participant group. The strongest association was detected for rs72704813 on chromosome 14q32.2 (OR = 0.64, CI 0.58-0.71, *P* = 4.0 × 10^-17^; Extended Data Table 4), located approximately 60 kilobases (kb) upstream of the *BDKRB2* gene. This variant has been identified as an expression quantitative trait locus (eQTL) for *BDRKB2* in the putamen (basal ganglia) and lung as well as for the *BDKRB1* gene in the putamen. Similar results were observed in participants with >5% 1KG-AFR-like genetic similarity (Extended Data Fig. 3 and Extended Data Table 6). In the self-identified White participant group, 77 SNVs from three loci reached genome-wide significance. The most significant association was observed for rs66515514 on chromosome 14q32.2 (OR = 0.72, CI 0.67-0.78, *P* = 4.5 × 10^-16^; Extended Data Table 5), located around 70 kb upstream of *BDKRB2.* Similar to rs72704813, this variant has been identified as an eQTL for *BDKRB2* in the putamen and lung as well as for *BDKRB1* in the putamen. Additional genome-wide significant associations included rs26209 on chromosome 5p15.2 (OR = 1.23, CI 1.16-1.31, *P* = 1.7 × 10^-11^; Extended Data Table 5), an eQTL for the *OTULINL* gene in multiple tissues (tibial nerve, esophagus muscularis, subcutaneous adipose, breast, visceral adipose, skeletal muscle, and tibial artery), and rs2903993 on chromosome 15q25.1 (OR = 3.97, CI 2.47-6.39, *P* = 1.2 × 10^-8^; Extended Data Table 5), located 25 kb downstream of the *CRABP1* gene. When results were meta-analyzed across the racial/ethnic groups (seven cohort-groups total), a total of 105 variants across two loci were identified at the genome-wide significant level (Extended Data Fig. 4). The most significant association was observed for rs72704813 on chromosome 14q32.2 (OR = 0.70, CI 0.66-0.74, *P* = 6.0 × 10^-31^). Additionally, genome-wide significance was observed for rs6866243 on chromosome 5p15.2 (OR = 0.84, CI 0.80-0.87, *P* = 2.6 × 10^-15^), an eQTL for the *OTULINL* gene in multiple tissues (tibial nerve, subcutaneous adipose, esophagus muscularis, breast, skeletal muscle, gastroesophageal junction, tibial artery, visceral adipose, sun-exposed skin, and not sun-exposed skin). The variant near *CRABP1* (rs2903993) was no longer genome-wide significant in the full meta-analysis (OR = 3.32, CI 2.13-5.18, *P* = 1.2 × 10^-7^).

**Figure 2.**
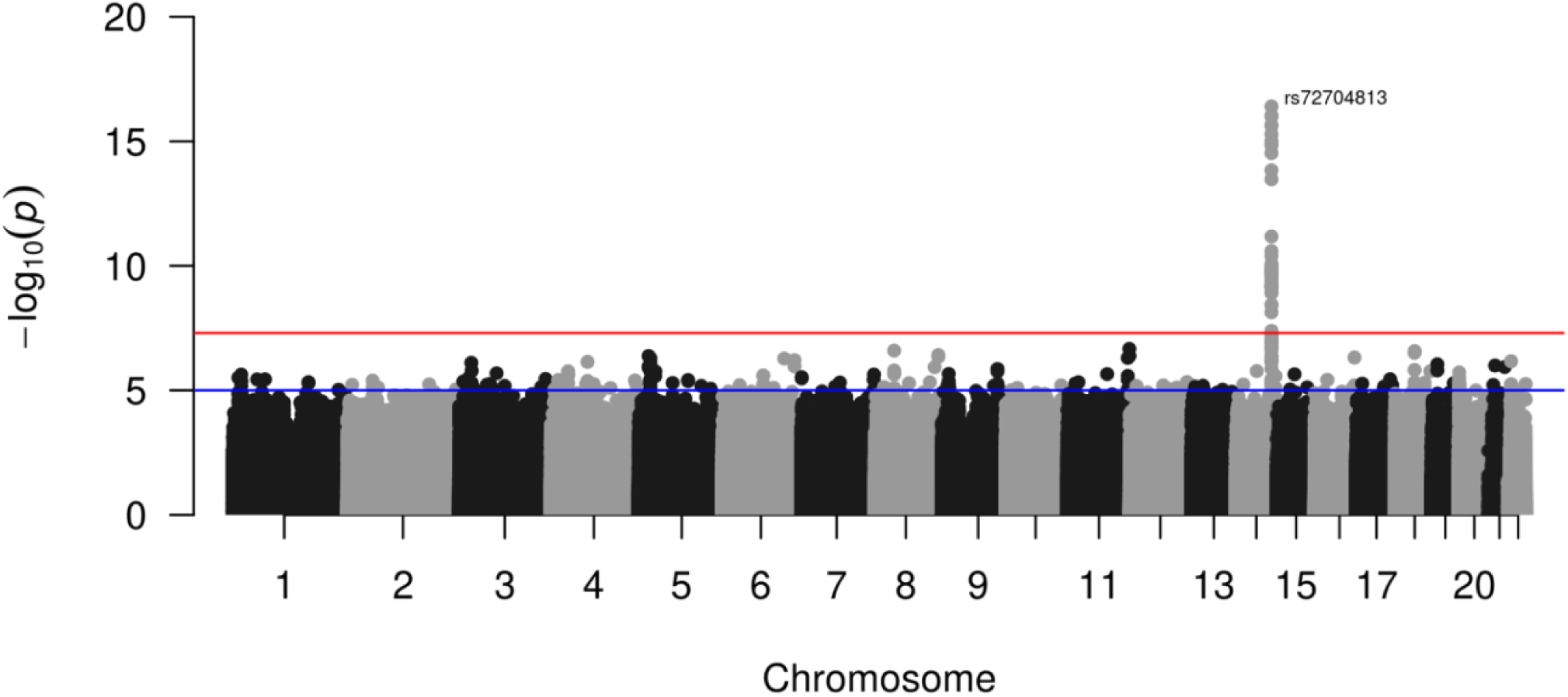
Meta-GWAS for RAAS inhibitor-induced angioedema in the Black participant group. (Case: N = 1657; Control: N = 16 567) GWAS, genome-wide association study; RAAS, renin-angiotensin-aldosterone system. The −log_10_ association *P* values (vertical axis) obtained from a fixed-effects meta-analysis of GWAS summary statistics in the Black participant groups across the All of Us Research Program (AoURP), Genetic Epidemiology Research in Adult Health and Aging (GERA), and Million Veteran Program (MVP) cohorts against their genomic positions (horizontal axis) are displayed. Significant associations were identified for variants near *BDKRB2*. The blue horizontal line indicates the prespecified threshold for a suggestive association (*P* = 1 × 10^-5^), and the red horizontal line indicates the threshold for genome-wide significance (*P* = 5 × 10^-8^).

**Figure 3.**
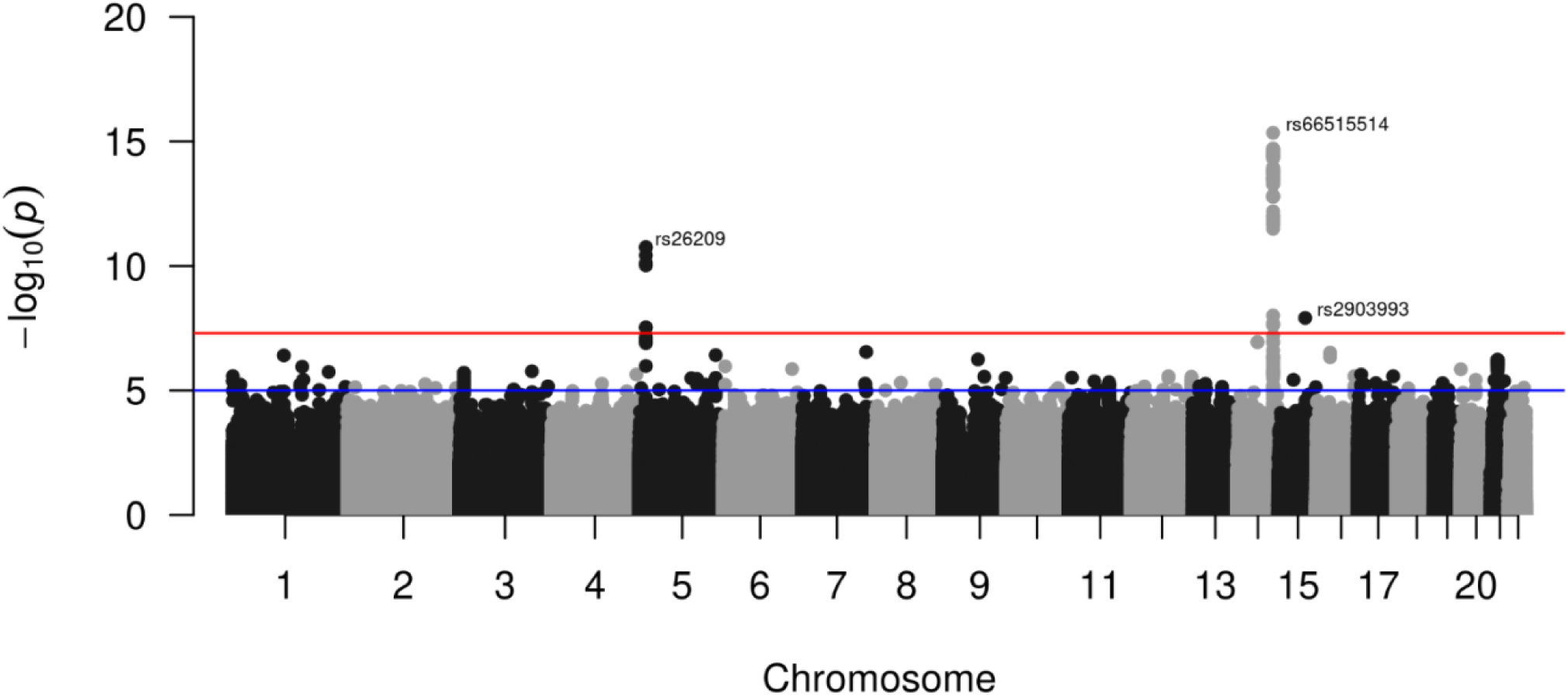
Meta-GWAS for RAAS inhibitor-induced angioedema in the White participant group. (Case: N = 2565; Control: N = 25 647) GWAS, genome-wide association study; RAAS, renin-angiotensin-aldosterone system. The −log_10_ association *P* values (vertical axis) obtained from a fixed-effects meta-analysis of GWAS summary statistics in the White participant groups across All of Us Research Program (AoURP), Genetic Epidemiology Research in Adult Health and Aging (GERA), and Million Veteran Program (MVP) cohorts against their genomic positions (horizontal axis) are displayed. Significant associations were identified for variants near *BDKRB2, OTULINL*, and *CRABP1.* The blue horizontal line indicates the prespecified threshold for a suggestive association (*P* = 1 × 10^-5^), and the red horizontal line indicates the threshold for genome-wide significance (*P* = 5 × 10^-8^).

### Heterogeneity of Genome-Wide Effects on RAAS Inhibitor-Induced Angioedema by Race

We identified four SNVs from three loci with significantly different effect sizes between Black and White participant groups. Fig. 4 displays the Manhattan plot of the heterogeneity tests, and Table 3 highlights the lead SNVs from each locus. The most significant association was observed for rs4905449 among lead SNVs, located on chromosome 14q32.2 (*P* = 1.1 × 10^-8^), approximately 55 kb upstream of the *BDKRB2* gene. This variant has been identified as an eQTL for *BDKRB2* in the lung, putamen, and cortex, as well as for the *BDKRB1* gene in the putamen. This association was followed by two intronic variants: rs12343822 on chromosome 9q21.2 (*P* = 2.0 × 10^-8^) within the *PSAT1* gene, and rs11943382 on chromosome 4q21.21 within the *LINC00989* gene (*P* = 4.0 × 10^-8^).

**Figure 4.**
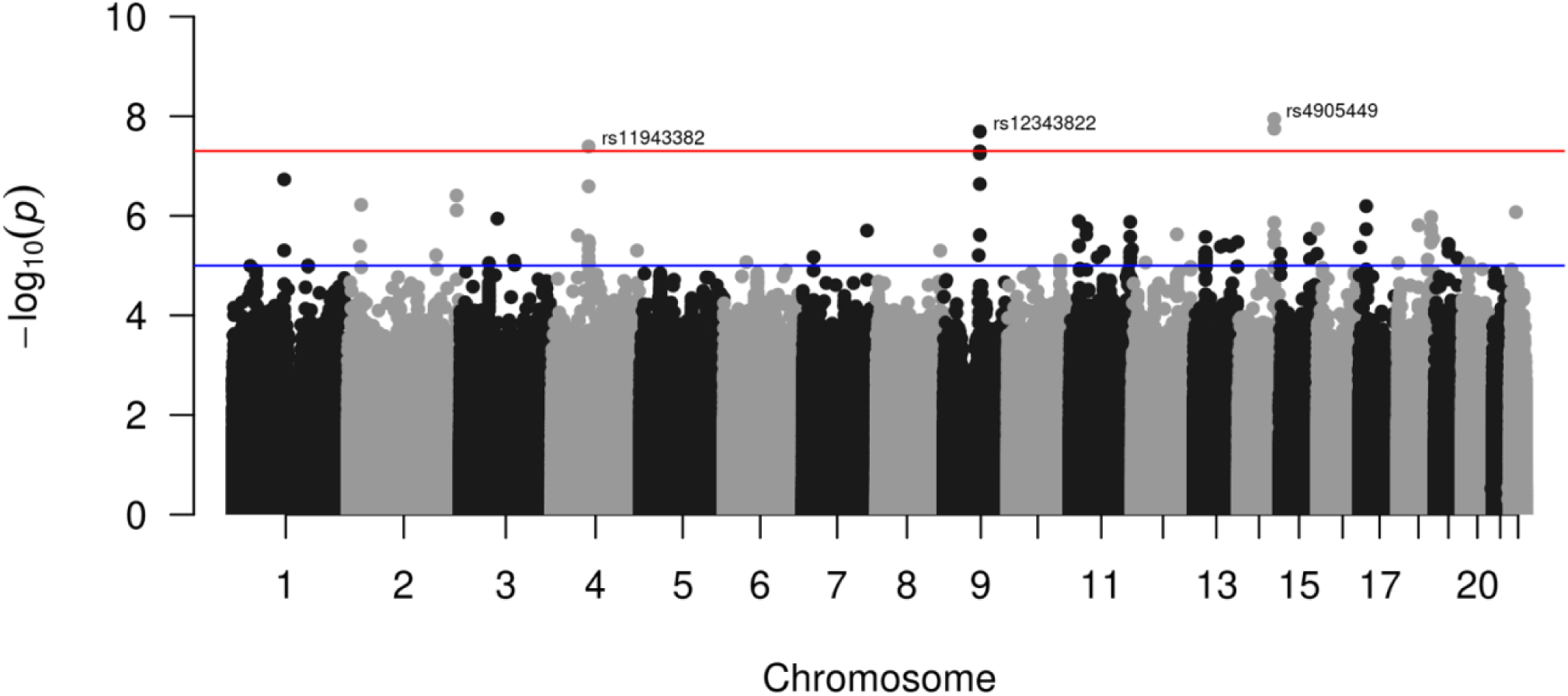
Genome-wide heterogeneity test for RAAS inhibitor-induced angioedema. RAAS, renin-angiotensin-aldosterone system. The −log_10_ association *P* values (vertical axis) obtained from genome-wide heterogeneity tests between the Black and White participant groups for genetic variants against their genomic positions (horizontal axis) are displayed. Significant associations were identified for variants near *BDKRB2.* The blue horizontal line indicates the prespecified threshold for a suggestive association (*P* = 1×10^-5^), and the red horizontal line indicates the threshold for genome-wide significance (*P* = 5 × 10^-8^).

**Table 3.**
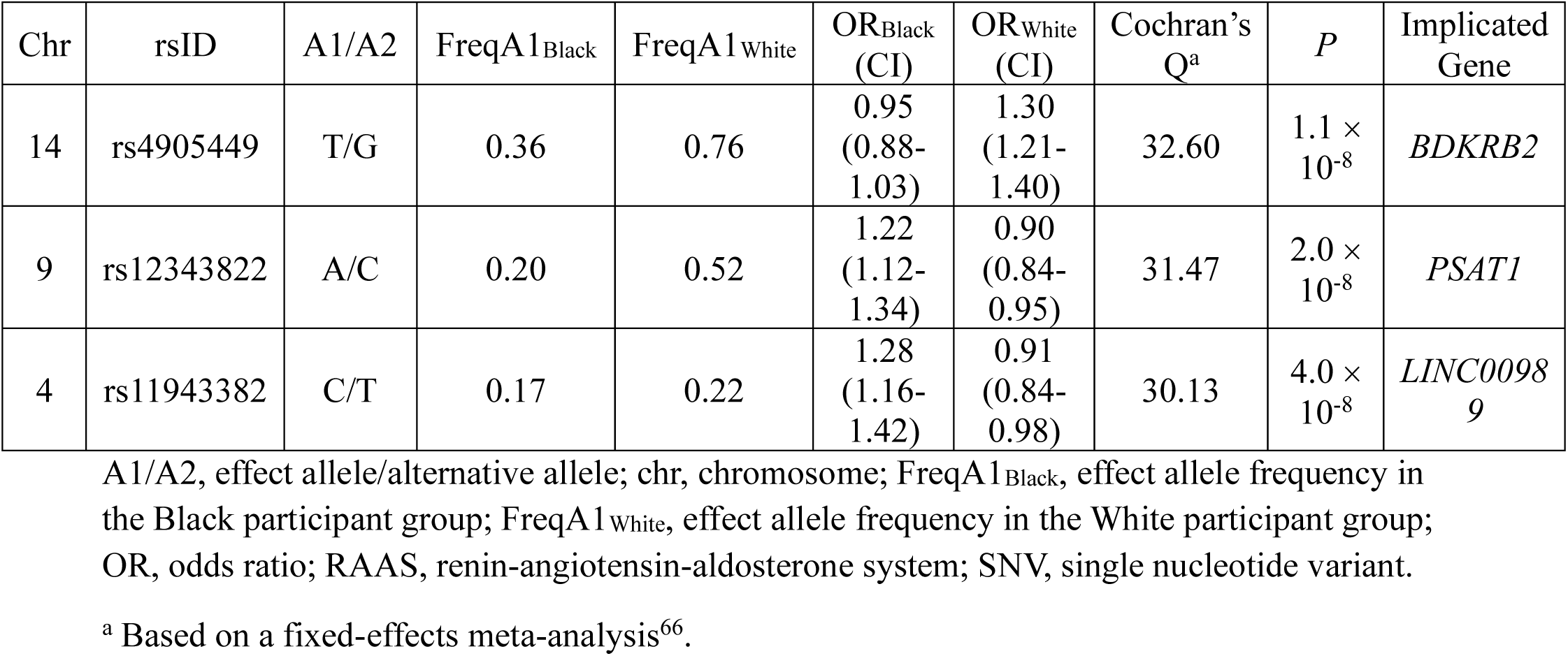
Significant lead SNVs in heterogeneity analyses of differential RAAS inhibitor-induced angioedema risk between the Black and White participant groups.

### Polygenic Factors Underlying Racially Different RAAS Inhibitor-Induced Angioedema Prevalence

Polygenic scores were constructed using summary statistics from (1) a Black versus White race heterogeneity test (PS_HET_) as well as (2) a GWAS meta-analysis for RAAS inhibitor-induced angioedema of three race/ethnicity groups (PRS_RAAS_), both using MVP data only. Sub-genome-wide variants were well-represented for both scores (76 597 and 834 variants for PRS_RAAS_ and PS_HET_, respectively). The two scores were only weakly correlated in an independent study population (Pearson *r* = −0.35, *P* < .001 in AoURP), supporting limited dependence. PRS_RAAS_ was associated with RAAS inhibitor-induced angioedema (OR = 1.57 per standard deviation, *P* = 2.1 × 10^-24^ in AoURP). The association of Black race with RAAS inhibitor-induced angioedema risk (OR = 4.01, *P* = 3.2 × 10^-67^ in AoURP) was partially attenuated when PS_HET_ was included in the model (OR = 2.66, *P* = 2.0 × 10^-17^ in AoURP; Fig. 5). Similarly, PRS_RAAS_ also partially attenuated the association of Black race with angioedema risk (OR = 2.39, *P* = 1.4 × 10^-18^ in AoURP; Fig. 5). When both PS_HET_ and PRS_RAAS_ were incorporated in the model together, the Black race association was further attenuated (OR = 1.80, P = 9.3 × 10^-7^ in AoURP; Fig. 5) compared to when either alone was added. PRS_RAAS_ had the strongest association with RAAS inhibitor-induced angioedema (OR = 1.51 per standard deviation, *P* = 1.4 × 10^-18^ in AoURP) when 1KG-AFR-like genetic similarity, SDOH, PS_HET_, and PRS_RAAS_ were all included in the model.

**Figure 5.**
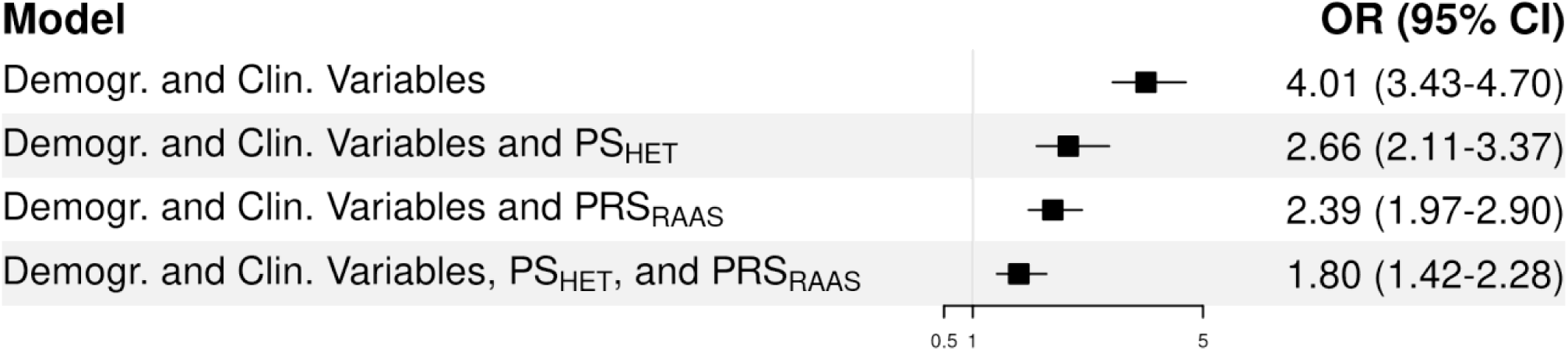
Association of Black race with RAAS inhibitor-induced angioedema risk in the AoURP cohort following polygenic score incorporation across multivariable logistic regression models AoURP, All of Us Research Program; clin., clinical; demogr., demographic; MVP, Million Veteran Program; PRS_RAAS_, RAAS inhibitor-induced angioedema polygenic risk score; PS_HET_, heterogeneity polygenic score; RAAS, renin-angiotensin-aldosterone system. The inclusion of PS_HET_ and PRS_RAAS_ (generated from MVP data) independently attenuated the association of Black race with RAAS inhibitor-induced angioedema risk in the AoURP cohort. When both PS_HET_ and PRS_RAAS_ were incorporated in the model simultaneously, the effect of Black race was further reduced.

**Figure 6.**
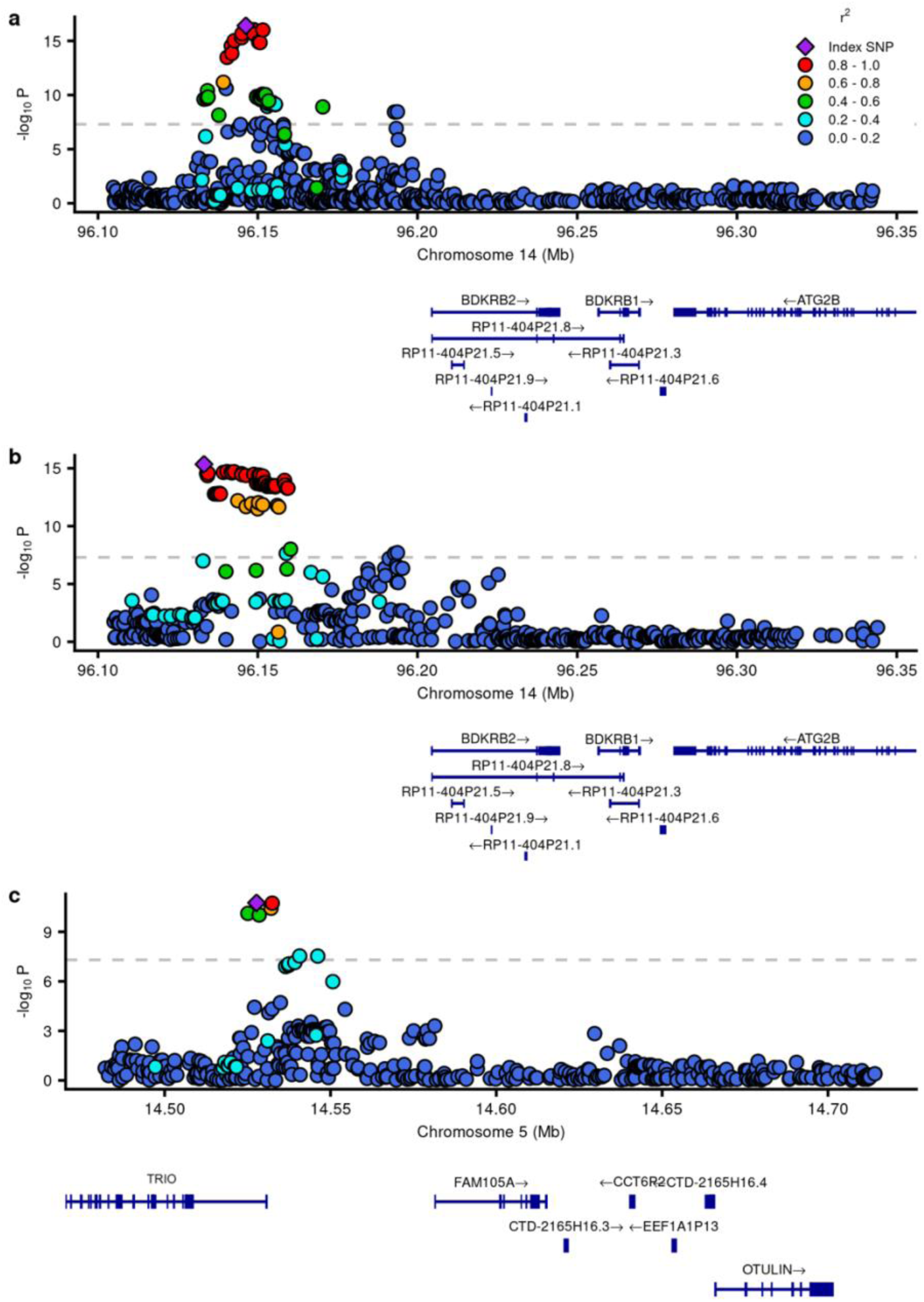
Zoom locus plot of loci reaching genome-wide significance for cross-race meta-GWAS of RAAS inhibitor-induced angioedema. GWAS, genome-wide association study; RAAS, renin-angiotensin-aldosterone system. The plots display the −log_10_ association *P* values (vertical axis) obtained from a fixed effects, cross-race meta-analysis of GWAS summary statistics across All of Us Research Program (AoURP), Genetic Epidemiology Research in Adult Health and Aging (GERA), and Million Veteran Program (MVP) cohorts against their genomic positions (horizontal axis). Results are shown for the *BDKRB2* locus in the Black participant group (**a**), the *BDKRB2* locus in the White participant group (**b**), and the *OTULINL* locus (displayed as *FAM105A*) in the White participant group (**c**) . The gray dotted horizontal line indicates the threshold for genome-wide significance (*P* = 5 × 10^-8^).

## Discussion

This study is the first to implicate *OTULINL* and *CRABP1* as novel loci of RAAS inhibitor-induced angioedema and to identify genome-wide significant (*P* < 5 × 10^−8^) variants of this ADR in self-identified Black participants. We are also the first to consider SDOH for RAAS inhibitor-induced angioedema, elucidating how they explain the differential prevalence of this ADR between Black and White participants relative to genetic determinants. Finally, we report initial evidence demonstrating how effect size and allele frequency variation may explain this differential ADR prevalence. Several strengths of this study enabled these novel findings. First, this study had 4461 cases, surpassing the previous largest study for RAAS inhibitor-induced angioedema, which included 1123 cases^15^. Furthermore, our study included 1657 cases in the Black participant group, far exceeding the only other GWAS in a similar study population, which included 202 cases^17^. Evaluating the full spectrum of RAAS inhibitors, rather than focusing solely on ACE inhibitors, also contributed to the increased sample size. Second, we leveraged SDOH from self-administered surveys, data not traditionally available in EHRs. Third, we employed a novel genome-wide heterogeneity test in our study design, which facilitated the identification of additional contributors to differential ADR prevalence between our racial groups.

We observed an increased risk of RAAS inhibitor-induced angioedema in Black participants compared to those of other racial backgrounds (as high as 5.6-fold increased risk when adjusting for potential confounding variables). This observation aligns with previous evidence, which has reported a twofold to fivefold increased risk in Black patients^5–7^. Notably, our observed fold increases fall on the higher end of this range. Additionally, we were generally able to reproduce the effect of previously established clinical and demographic variables for this drug response phenotype (including higher risk associated with ACE inhibitors compared with ARBs^7^, female sex^5,23^, advanced age^24–26^, and Hispanic ethnicity^26^). Altogether, these findings that replicate prior clinical observations support our phenotype’s accuracy as we proceeded to interrogate genetic and social determinants of response.

At the genome-wide significant level, we identified variants implicated for *BDKRB2*, *OTULINL*, and *CRABP1* as predictors of RAAS inhibitor-induced angioedema. Among these, *BDKRB2* has been previously identified for this drug response phenotype through GWASs^15,16^. *BDKRB2* encodes the bradykinin receptor B2, which binds bradykinin. Bradykinin is a key mediator of vascular permeability and inflammation, and its accumulation following ACE inhibition may contribute to angioedema development^27^. The *OTULINL* gene is paralogous to the *OTULIN* gene^28^, which encodes a protein crucial for regulating inflammation and plays a role in OTULIN-related autoinflammatory syndrome (ORAS) pathology^29,30^. Although *OTULINL* encodes an enzymatically inactive protein, it may mediate protein-protein interactions within inflammatory signaling pathways^28^. Additionally, given the proximity of *OTULINL* and *OTULIN*, it is plausible that the identified lead SNV in this locus tags a causal variant affecting *OTULIN*. Interestingly, although ORAS and angioedema represent clinically distinct entities, both conditions share common features of subcutaneous tissue inflammation and swelling. This finding could theoretically have arisen from clinical misdiagnosis of ORAS as angioedema. However, due to the rarity of ORAS and the typical timing of diagnosis (early infancy)^31^, the likelihood of this is extremely low. *CRABP1* has also been reported to play a protective role against inflammation^32^, suggesting that perhaps its dysregulation may contribute to angioedema susceptibility. Future mechanistic studies are warranted to determine the precise role of *OTULINL/OTULIN* and *CRABP1* in RAAS inhibitor-induced angioedema.

Importantly, this study identified a genome-wide association for *BDKRB2* in self-identified Black participants, who predominantly exhibit 1KG-AFR-like genetic similarity. This finding is notable, as most genomic studies are conducted in Eurocentrically biased study populations, producing results that can be generalized to only a fraction of the globe^19^. For example, fewer than five pharmacogenomic studies to date have identified variants meeting the threshold of genomic significance in participants with predominantly 1KG-AFR-like genetic similarity. This situation can further worsen preexisting health disparities, due to unrecognized pharmacogenetic risk alleles enriched in groups genetically dissimilar to those that are 1KG-EUR-like^14^. These findings overall demonstrate the strong importance of bradykinin in RAAS inhibitor-induced angioedema for participants of 1KG-AFR-like genetic similarity, which strengthens our confidence in this pathway as a more generalizable contributor across groups. These findings help set the stage for future precision medicine health equity work that leverages historically underrepresented study populations to advance pharmacogenetics. Efforts are underway^33,34^.

We identified increased alcohol consumption, lower education level, higher income, not being married, less walking, better general quality of life, less general satisfaction with social activities and relationships, better general social role performance, and longer years lived at the current address as SDOH associated with increased RAAS inhibitor-induced angioedema risk. Among these, alcohol consumption had the strongest association. In the GERA cohort, we found that an extra alcoholic beverage daily increased the risk of this ADR by approximately 30%. Alcohol is known to have numerous acute effects. Fluid retention, mast cell degranulation, bradykinin signaling, urticaria, and complement activation, in particular, are alcohol effects that may be lowering the threshold for RAAS inhibitor-induced angioedema^35–39^. However, future studies are necessary to validate these findings. Additionally, the inclusion of more contemporary SDOH (e.g., discrimination, perceived stress) and their interactive effects on RAAS inhibitor-induced angioedema would enable a more comprehensive investigation.

A critical objective of the current study was to elucidate factors explaining the increased RAAS inhibitor-induced angioedema risk in Black patients that have been observed clinically for decades. Even the strongest aforementioned genetic and social predictors of this ADR may not necessarily contribute to this differential response (e.g., if they have similar prevalence and effect sizes between groups). Our study reveals multiple findings regarding these differing drug effects across groups. First, we discovered that while 1KG-AFR-like genetic similarity completely abrogated the racial effect on angioedema, SDOH had no impact. This implicates genetic determinants. Second, close inspection of our genome-wide significant associations suggests that these variants alone could not possibly explain (and possibly contradict) angioedema differences between groups (Table 4); this finding strongly points to a polygenic architecture underlying differences. Third, polygenic scores attenuated the statistical effect of 1KG-AFR on race, suggesting that specific variants rather than broad genetic similarity are a more precise predictor. Fourth, PRS_RAAS_ was the strongest predictor supporting a role for differing allele frequencies in explaining differential effects by racial groups. Fifth, our genome-wide heterogeneity test results (including the impact of incorporating PS_HET_ into our models) indicate that incongruent genetic effect sizes also play a role orthogonal to allele frequency differences. In particular, lead heterogeneity SNV rs4905449 (of *BDKRB2*) was not in linkage disequilibrium with independent associations from the *BDKRB2* locus for the RAAS inhibitor-induced angioedema GWAS of the Black participant group (*r*^2^ = 0.10 and 0.01, for rs72704813 and rs881163, respectively).

**Table 4.** Contribution of SNVs to increased risk of RAAS inhibitor-induced angioedema in self-identified Black compared to White participant groups based on frequency or effect.

| rsID<sup>a</sup> | Locus | Chr | Study<sup>b</sup> | A1<sup>c</sup> | By FreqA1?<sup>d</sup> | By EffectA1?<sup>e</sup> |
| --- | --- | --- | --- | --- | --- | --- |
| rs72704813 | 14q32.2 | 14 | Black | G | Yes | Yes |
| rs66515514 | 14q32.2 | 14 | White | C | No | No |
| rs26209 | 5p15.2 | 5 | White | G | No | No |
| rs2903993 | 15q25.1 | 15 | White | G | Yes | NA |
| rs4905449 | 14q32.2 | 14 | Heterogeneity | T | No | No |
| rs12343822 | 9q21.2 | 9 | Heterogeneity | A | No | Yes |
| rs11943382 | 4q21.21 | 4 | Heterogeneity | C | No | Yes |

chr, chromosome; GWAS, genome-wide association study; NA, not applicable; RAAS, renin-angiotensin-aldosterone system; SNV, single nucleotide variant.
<sup>a</sup> Genome-wide significant lead SNVs from three studies of RAAS inhibitor-induced angioedema: a meta-GWAS of Black participants across three cohorts, a meta-GWAS of White participants across the same three cohorts, and a genome-wide heterogeneity study comparing genetic effects from the two aforementioned summary statistics. The three cohorts are All of Us Research Program (AoURP), Genetic Epidemiology Research in Adult Health and Aging (GERA), and Million Veteran Program (MVP).
<sup>b</sup> The specific study where each variant was identified as a lead significant SNV. *Black*, *White*, and *Heterogeneity* correspond to the meta-GWAS of RAAS inhibitor-induced angioedema in Black and White participant groups and the heterogeneity analysis of differential angioedema risk between these groups, respectively.
<sup>c</sup> The allele associated with increased risk of RAAS inhibitor-induced angioedema (i.e., the risk allele). For SNVs where risk allele designation varies by racial group (i.e., SNVs from the Heterogeneity analysis), the allele with the larger absolute beta value was chosen.
<sup>d</sup> *By FreqA1?* refers to whether the frequency of A1 (risk allele) is greater in the Black participant group compared to the White participant group. *Yes* indicates that the variant contributes to increased risk of RAAS inhibitor-induced angioedema in Black participants based on frequency differences.
<sup>e</sup> *By EffectA1?* refers to whether the odds ratio for A1 (risk allele) is higher in the Black participant group compared to the White participant group. *Yes* indicates that the variant contributes to increased risk of RAAS inhibitor-induced angioedema in Black participants based on differences in effect. *NA* indicates that rs2903993 was not available in the meta-analysis for the Black participant group, as data for this variant were only accessible in the GERA cohort.

Differing genetic effects were also found at *PSAT1* and *LINC00989* loci, which in contrast to *BDKRB2* were distinct from those identified through our within-group and across-group GWASs. *PSAT1* is a key enzyme in the serine synthesis pathway^40^. The extracellular serine levels have been shown to be crucial for suppressing aberrant cytokine production^41^, suggesting that its dysregulation may contribute to angioedema susceptibility. *LINC00989* has been implicated in pericyte-mediated fibrosis on cardiac fibroblasts, suggesting its potential role in heart failure progression^42^. Prior studies have demonstrated an elevated risk of RAAS inhibitor-induced angioedema in those with chronic heart failure^5^. This association may thus reflect differences in the racial prevalence of chronic heart failure. More studies are necessary to better understand the reasons generally for varying effect sizes. Differing linkage disequilibrium patterns and gene-by-environment effects may play a large role^43^. Altogether, these findings implicate genetic determinants enriched within genomes similar to 1KG-AFR as the primary drivers of differential angioedema drug response by race. These findings also demonstrate that heterogeneity analyses can capture additional genetic contributions across study groups that traditional GWASs may miss.

The findings presented have important clinical implications. Compared to earlier heart failure treatment guidelines^9–11^, the most recent version has removed the mention of differential RAAS inhibitor-induced angioedema by race^12^. This is unrelated to new evidence and without any attempts to address the underlying health disparity; it likely represents a reaction to recent calls for removing race from medicine^13^. In contrast, our study facilitates the removal of race from guidelines through an evidence-based approach. We show that genetic variants, rather than race in and of itself, explain the differential effects. These findings suggest that a clinical tool may more precisely predict for whom RAAS inhibitors should be avoided (to prevent angioedema). For these patients, depending on the indication, there are alternatives that can be added to their pharmacotherapeutic regimen (e.g., sodium-glucose cotransporter-2 inhibitors for heart failure). This clinical tool, if applied universally, could address this disparity by directly reducing the elevated rates observed in Black patients, rather than simply ignoring it. Before implementation, however, further studies are necessary to refine this tool and establish its clinical validity. In addition, although SDOH did not explain racial differences in response, we did find them to impact risk overall. Thus, our findings implicate the potential role of addressing social factors for RAAS inhibitor-induced angioedema in routine care as well. Interventions that address social stressors are increasingly being recommended by clinical guidelines for cardiovascular health^12^.

This study is not without limitations. First, causality cannot be definitively inferred for our strongest genetic associations. Nevertheless, this is the largest study of its kind for study populations of 1KG-AFR-like genetic similarity and the first to demonstrate genome-wide significance in these participants. Therefore, findings presented here provide biological insight and establishes a strong foundation for future studies that perform functional validation of our associations. Second, our findings cannot be generalized to certain patient groups who were not well represented in our study (e.g., participants with genetic similarity to the 1KG East Asian continental superpopulation). However, our research objectives and hypotheses originated from a well-established health disparity observed in self-identified Black patient groups. Moreover, extremely small sample sizes precluded investigations into these study populations as results would be uninterpretable. Additionally, pooling small groups in an attempt to be inclusive does not actually increase inclusivity since results will reflect the majority population^44^. Future studies with larger sample sizes enabling population-specific analyses for underrepresented groups are needed.

In summary, this study has identified genetic and social determinants that increase the risk of RAAS inhibitor-induced angioedema overall as well as between racial groups. Clinically, this research provides evidence supporting the removal of race and the implementation of precision medicine in treatment guidelines to optimize drug safety for all patient groups taking RAAS inhibitors. Future studies should prioritize (1) clinical validation, (2) additional social and genetic factors, and (3) clinical translation.

## Data availability

Raw data, including individual-level data from GERA and AoURP, can be accessed by investigators through an application and review process. MVP data are accessible to all Veterans Affairs-affiliated researchers through an application process. Summary-level data are available upon written request.

## Code availability

Details of the SAIGE GWAS pipeline can be found at https://saigegit.github.io/SAIGE-doc/, and the meta-GWAS implementation in PLINK is described at https://zzz.bwh.harvard.edu/plink/metaanal.shtml. Polygenic score construction was performed following the PRSice-2 pipeline (https://choishingwan.github.io/PRSice/).

## Methods

### Data Resources

This study was conducted leveraging data from three independent EHR-linked biobanks. Institutional Review Board (IRB) approval was obtained from both Kaiser Permanente and the University of California. Participants gave written informed consent.

Kaiser Permanente (KP) is an integrated health system providing comprehensive healthcare to approximately 12.5 million members across eight regions in the United States: Colorado, Georgia, Hawai’i, Mid-Atlantic, Northern California, Northwest, Southern California, and Washington^45^. The GERA cohort consists of around 100 thousand KP Northern California Region Medical Care Plan members with self-administered survey data on demographic, behavioral, social, and environmental factors as well as genome-wide genotyping data linked to EHRs. Participants range in age from 18 to 100 years, with the majority self-identifying as non-Hispanic White (81%). Among the remaining participants, over 3 thousand individuals self-identified as Black^22^. The EHR data included pharmacy (dispensing) records and diagnostic codes. Genotyping was performed using four customized Affymetrix Axiom arrays designed to maximize genome-wide coverage^46^ followed by quality control procedures^47^. Genomic imputation followed the approach previously described^48^ using the 1KG Project as the reference panel with pre-phasing in SHAPEIT v2.r727 and imputation in IMPUTE2 v2.3.0.

The National Institutes of Health (NIH) AoURP is an effort aimed at building a comprehensive data repository integrating health questionnaires, EHRs, physical measurements, and biospecimens, with the goal of enrolling a diverse cohort of more than one million adults in the United States^21^. For this study, the Curated Data Repository version 8 (within the Controlled Tier dataset, the only Tier that genomic data can be accessed) was leveraged, which included EHR data from approximately 400 thousand participants, survey data from more than 600 thousand participants, and whole genome sequencing (WGS) data from more than 400 thousand participants^49^. The EHR data included drug exposures (documenting medication utilization) and condition occurrences (records indicating the presence of diseases or medical conditions), both of which are standardized using the Observational Medical Outcomes Partnership Common Data Model^21^. WGS data generated by the AoURP Genomics Investigators were available either as short-read or long-read sequencing^21^. For this study, we utilized three short-read callsets: Allele Count/Allele Frequency (ACAF) Threshold, Exome, and ClinVar. Additionally, SDOH were available in the AoURP cohort from self-administered survey data.

The Veterans Health Administration (VHA) is a large, integrated health system serving over 9 million enrolled Veterans across the United States^50^. For this study, we accessed EHRs from the Corporate Data Warehouse of the VHA, a repository of clinical data^51^. The EHR data included patient demographics, pharmacy records, and diagnostic codes. The MVP is a biobank linked to VHA EHRs comprising approximately 650 thousand participants with genotyping data available^52^. The majority of MVP participants self-identified as White (74%), followed by Black (18%)^53^. Genotyping was performed using a customized Affymetrix Axiom array followed by quality control. Genomic imputation was conducted using the 1KG phase 3 v5 reference panel with EAGLE v2.4 and Minimac4^54^.

### Study Populations and RAAS Inhibitor-induced Angioedema Drug Response

The following classes of RAAS inhibitors were eligible for inclusion in our analyses: ACE inhibitors, ARBs, and direct renin inhibitors (DRIs). We included all RAAS inhibitor users; although the strongest effects for angioedema have been observed with ACE inhibitors, there is evidence suggesting that ARBs and DRIs also elevate risk^55^. RAAS inhibitor users, defined as participants who were ever-users of these therapeutic classes, were identified within each cohort using pharmacy records.

We generated a drug response phenotype using a case-control study design. In the GERA and MVP cohort, RAAS inhibitor-induced angioedema cases were defined as RAAS inhibitor users with a diagnosis record of angioedema (International Classification of Diseases [ICD]-9 code 995.1^56^ or ICD-10 code T78.3XXA^57^). In the AoURP cohort, RAAS inhibitor-induced angioedema cases were defined as RAAS inhibitor users with a report of angioedema in the drug exposure, observation, or condition occurrence domain (standard concept names: “angioedema,” “angioedema of lip,” “ACE inhibitor-aggravated angioedema,” “angioedema due to angiotensin-converting-enzyme inhibitor,” and “angioedema of eyelid”). In all cohorts, a diagnosis of angioedema was eligible as a case if it was within the timeframe of any RAAS inhibitor pharmacy record (from the start date of the record to up to 7 days following the end date of the same record^33^). For participants with multiple angioedema diagnoses, only the initial diagnosis was considered. If the initial diagnosis occurred outside the RAAS inhibitor therapy timeframe, the participant was deemed ineligible as a case and was excluded from the study population. To designate the index record, the GERA cohort used the earliest RAAS inhibitor record for which the angioedema diagnosis occurred within its timeframe, whereas the AoURP and MVP cohorts used the latest RAAS inhibitor record for which the angioedema diagnosis occurred within its timeframe.

The prevalence of RAAS inhibitor-induced angioedema in each cohort was calculated by dividing the number of identified RAAS inhibitor-induced angioedema cases by the total number of RAAS inhibitor users (excluding ineligible cases as described above).

RAAS inhibitor users with no record of angioedema met the criteria as potential controls. In all cohorts, only the earliest RAAS inhibitor pharmacy record was considered as a potential index record for each individual. A RAAS inhibitor pharmacy record was eligible as a potential index record for controls if (1) the pharmacy record start date was not identical to the start date of another RAAS inhibitor record and (2) no evidence of discontinuation was observed for that record. We defined evidence of discontinuation as the absence of a subsequent pharmacy record for the same drug type, with a gap of ≤6 months between the end date of the earlier record and the start date of the subsequent record^58^.

For all index pharmacy records (from cases and controls), the start date was deemed the index date, except for cases in the AoURP cohort, where the diagnosis date was used as the index date. The “full” study population consisted of all eligible cases and controls.

Since pharmacoepidemiology studies are particularly prone to unmeasured confounding between cases and controls (e.g., channeling bias, healthy user bias)^59^, we performed matching as a prespecified study design feature. Each case was matched to 10 controls based on the following criteria at the time of index date: sex, age (≤3-year difference in the GERA cohort; ≤5-year difference in the AoURP and MVP cohorts), and self-identified race. Additional matching criteria varied by cohort. In the GERA cohort, drug class, reagent kit used for genotyping^47^, and genomic imputation group were additionally included as matching criteria. In the AoURP and MVP cohorts, drug type was included as an additional criterion. Furthermore, in the GERA and AoURP cohorts, self-identified ethnicity (Hispanic versus non-Hispanic) and whether the participant exhibited >5% 1KG-AFR-like genetic similarity or not were included as matching criteria. Cases without 10 matched controls were deemed ineligible for further analyses. The “matched” study population consisted of all eligible cases and their corresponding matched controls (to account for potential unmeasured confounding within our observational cohorts). In the MVP cohort, some participants were excluded from further analyses due to withdrawal of informed consent.

### Justification for the Use of Population Descriptors

The integration of pharmacogenetics into clinical practice has been found to improve drug safety outcomes^60^. However, Eurocentrically biased study populations currently predominate findings in pharmacogenetic research, potentially exacerbating existing health disparities^19^. This study’s aforementioned objective falls in line with the ultimate goal of improved health outcomes for all patient groups eligible for RAAS inhibitor therapy, especially groups that remain understudied in pharmacogenetics.

After careful consideration, we determined that the use of population descriptors was crucial to conduct our health equity study. In particular, our analyses were conducted in and across the following groups (not all being mutually exclusive): self-identified Black participants, self-identified White participants, Hispanic/Latino participants, and participants with >5% 1KG-AFR-like genetic similarity. The first two study groups served as our primary analysis groups. We decided to use these racially defined groups because our study hypothesis and objective are based on well-established differential RAAS inhibitor-induced angioedema rates observed clinically between Black and White patients (considered a health disparity based on the commonly cited NIMHD definition^8^). As described above, to control for potential confounding, we also used race and ethnicity for matching (e.g., bias due to potential surveillance differences in angioedema by race). However, consistent with recent guidance^61^, these descriptors were not used as proxies for genetic similarity but rather to contextualize observed racial differences in drug safety outcomes as health disparities. In contrast, the >5% 1KG-AFR-like genetic similarity group was included as a sensitivity analysis group for the self-identified Black participant group to explore potential genetic contributions to angioedema susceptibility, under the hypothesis that 1KG-AFR-like genetic similarity-related variants play a role irrespective of self-identified race (i.e., genetic variants enriched on haplotypes associated with 1KG-AFR-like genetic similarity increase susceptibility). Unlike self-identified race and ethnicity, which reflect sociocultural and environmental factors, we used this categorization as a proxy for genetic similarity in this study.

### Statistical Analyses

Multivariable logistic regression was employed in the full study populations within each cohort to assess the association of clinical and demographic variables (age per year, female sex, drug type, Black race, and Hispanic ethnicity) with our RAAS inhibitor-induced angioedema drug response outcome.

SDOH and 1KG-AFR-like genetic similarity proportion were added to the above models for GERA and AoURP cohorts (due to data availability of SDOH in those cohorts only).

We ran logistic regression analyses within stratified racial groups in the full study populations of GERA and AoURP (with age, sex at birth, and drug class as covariates) to select SDOH for model inclusion above as follows. SDOH survey questions with a response rate of <80% were excluded from the analysis. For the remaining questions, rare answer categories (minor response frequencies <2%) were merged to ensure each had a minor response frequency >2%. In the GERA cohort, binary categories (either by design or as a result of merging) with minor response frequencies <5% were also excluded. For the purpose of data reduction, all questions related to diet (available only in GERA) were consolidated into two dietary pattern scores: a Western or “unhealthy” dietary pattern (characterized by higher consumption of meats and fried foods) and the prudent or “healthier” dietary pattern (characterized by greater intake of fruits and vegetables).^62^ Those SDOH associated with our drug response outcome in either of the stratified racial groups (α = 0.05) were carried forward into the full study population models for GERA and AoURP.

GWASs of autosomal variants for RAAS inhibitor-induced angioedema were performed within each analysis group (Black, White, >5% 1KG-AFR in each of the three cohorts, and Hispanic/Latino in the MVP cohort) using a logistic mixed model implemented in Scalable and Accurate Implementation of Generalized Mixed Model (SAIGE)^63^. An additive genetic effect was assumed, adjusted for matching variables and the top six eigenvectors for each group. An α = 5 × 10⁻⁸ was used to determine statistical significance^64^.

PLINK^65^ was used to conduct fixed-effect meta-analyses of the above GWAS summary statistics. Further details are available in the Supplementary Material.

To assess whether variants exhibit differential effects between Black and White participant groups for RAAS inhibitor-induced angioedema (i.e., a gene-environment interaction), a genome-wide heterogeneity test was conducted using Cochran’s Q test^66^ on race-conscious summary statistics across all three cohorts. An α = 5 × 10⁻^8^ was used to determine statistical significance.

Polygenic scores were constructed using summary statistics from the heterogeneity test (PS_HET_) and GWAS meta-analysis combining racial groups (PRS_RAAS_) based on MVP data only with PRSice-2^67^. As part of the quality control process, variants with minor allele frequencies (MAF) <0.01, imputation information scores <0.8, duplicate SNVs, and strand-ambiguous SNVs were removed from the base dataset. In the target dataset, variants with MAF <0.01, a genotyping rate <0.1, and a Hardy-Weinberg Equilibrium *P* value (calculated separately for each racial group) <1.0 × 10^-7^ were excluded. Linkage disequilibrium reference data were derived from 1KG-AFR phase 3. Variants for the polygenic scores were selected using *P* value thresholds ranging from 5.0 × 10^-8^ to .5, in increments of 5.0 × 10^-5^. The association between the polygenic scores and RAAS inhibitor-induced angioedema risk (odds) was then assessed by adding these variables to our original logistic regression model of the full AoURP study population (e.g., including clinical and demographic variables).

The above analyses were conducted using R software (R Foundation for Statistical Computing. Version 4.4.2. Available at https://www.R-project.org) or Python (Python Software Foundation. Version, 3.13.2. Available at https://www.python.org) unless otherwise indicated.

## Acknowledgements

Research reported in this publication was supported by the National Human Genome Research Institute of the National Institutes of Health under Award Number R01HG012824 and the Office of the Director - All of Us (OD-AoURP). The content is solely the responsibility of the authors and does not necessarily represent the official views of the National Institutes of Health or the OD-AoURP. We thank the Kaiser Permanente Northern California members who have generously agreed to participate in the Kaiser Permanente Genetic Epidemiology Research on Adult Health and Aging (GERA) cohort. The development of GERA was supported by grants from the Robert Wood Johnson Foundation, the Wayne and Gladys Valley Foundation, the Ellison Medical Foundation, and Kaiser Permanente Community Benefit Programs. This research is also based on data from MVP, Office of Research and Development, VHA. We thank the Veterans who generously agreed to participate in MVP. This work was supported using resources and facilities of the Veterans Affairs Informatics and Computing Infrastructure (VINCI), including data analytics conducted by its Precision Medicine research team, which is funded under the research priority to Put Veterans Affairs Data to Work for Veterans (VA ORD 24-D4V-02). Kathryn Pridgen assisted with the literature review. This article does not reflect the position or policy of the Department of Veterans Affairs or the United States government. Some data used for the analyses described in this manuscript were obtained from: the GTEx Portal (https://www.gtexportal.org/home/) on 10/23/25.

## Author information

These authors contributed equally: Nanase Toda, Tanushree Haldar, Craig C. Teerlink. These authors jointly supervised this work: Julie A. Lynch, Elad Ziv, Akinyemi Oni-Orisan. Contributions N.T. and A.O. wrote the manuscript. N.T., C.T., E.Z., and A.O. contributed to the design of the research. T.H., C.C.T., D.H., M.L., C.T., C.I., and J.A.L. contributed to the acquisition of the data. N.T., T.H., C.C.T., D.H., P.D., S.H., C.T., J.A.L, A.B., E.Z., and A.O. contributed to the analysis or interpretation of the data. All authors approved the submission of this manuscript.

## Ethics declarations

## Tables

**Extended Data Table 1.** Demographic and clinical characteristics in participants with >5% 1KG-AFR-like genetic similarity of the matched study population.

|  |  | GERA (N=616) | AoURP (N=3465) | MVP (N=15 435) |
| --- | --- | --- | --- | --- |
| Case count (n)<sup>a</sup> |  | 56 | 315 | 1419 |
| Control count (n)<sup>b</sup> |  | 560 | 3150 | 14 016 |
| Age, median (IQR)<sup>c</sup> |  | 65.5 (11.5) | 57.8 (13.8) | 60.6 (11.2) |
| Sex<br>(n, %) | Male | 308 (50) | 1232 (36) | 13 899 (90) |
|  | Female | 308 (50) | 2233 (64) | 1436 (10) |
| Drug<br>class<br>(n, %) | ACE<br>inhibitors | 561 (91) | 3133 (90) | 14 315 (93) |
|  | ARBs | 55 (9) | 352 (10) | 1120 (7) |
| Hispanic ethnicity<br>(n, %) |  | 77 (12) | 341 (10) | 450 (3) |

1KG-AFR, 1000 Genomes African superpopulation; ACE, angiotensin-converting enzyme; AoURP, All of Us Research Program; ARBs, angiotensin-receptor blockers; GERA, Genetic Epidemiology Research in Adult Health and Aging; IQR, interquartile range; MVP, Million Veteran Program.
<sup>a</sup> Case count refers to the number of ever-users of renin-angiotensin-aldosterone (RAAS) inhibitors with a diagnosis record of angioedema within the eligible timeframe.
<sup>b</sup> Control count refers to the number of ever-users of RAAS inhibitors without a record of angioedema, whose initial RAAS inhibitor record met the inclusion criteria and who were matched to cases in a 1:10 ratio. In the MVP cohort, some participants were excluded due to withdrawal of their informed consent.
<sup>c</sup> Age corresponds to the participant's age at the start date of their index pharmacy record, defined as the eligible RAAS inhibitor record used for subsequent analyses.

**Extended Data Table 2.**
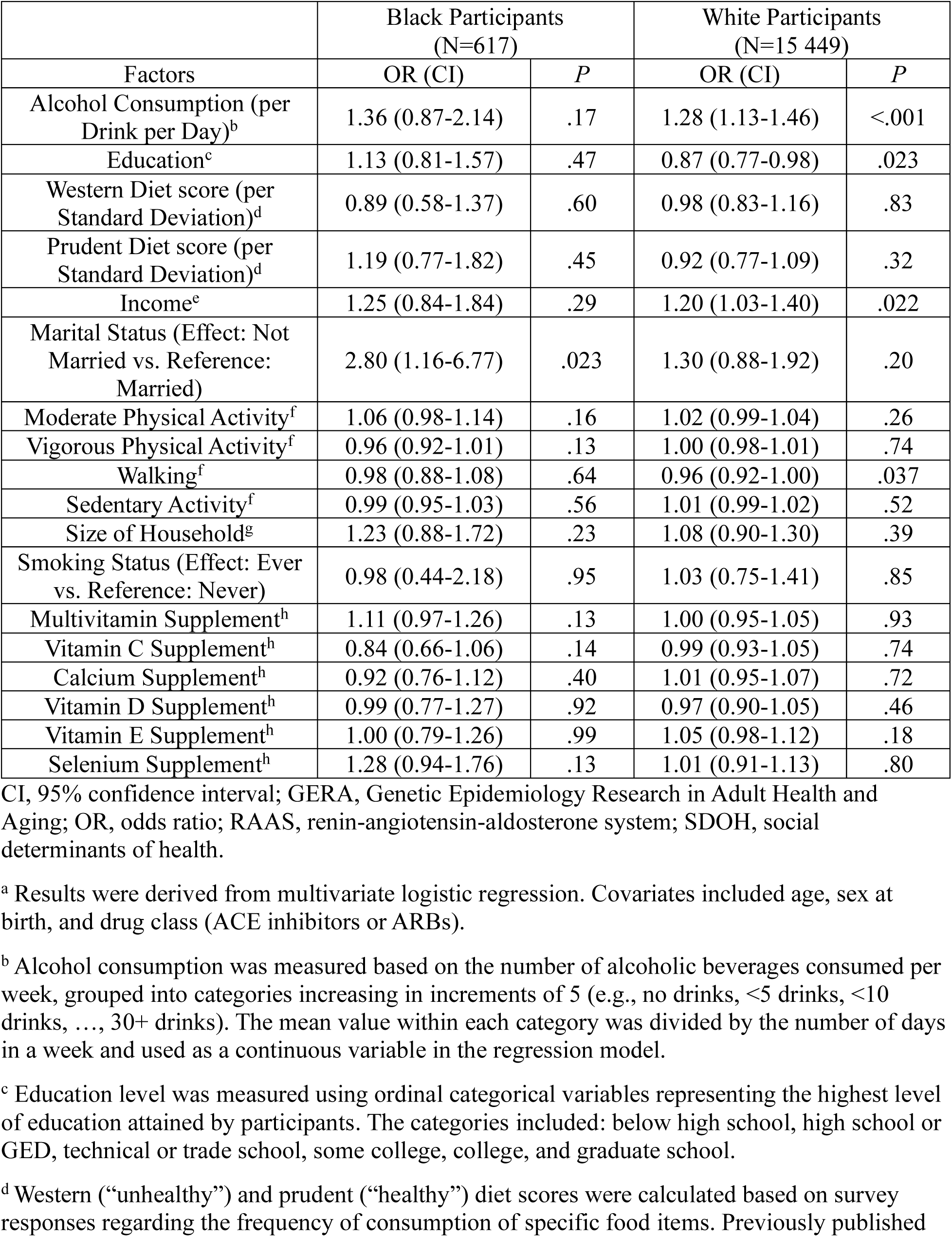
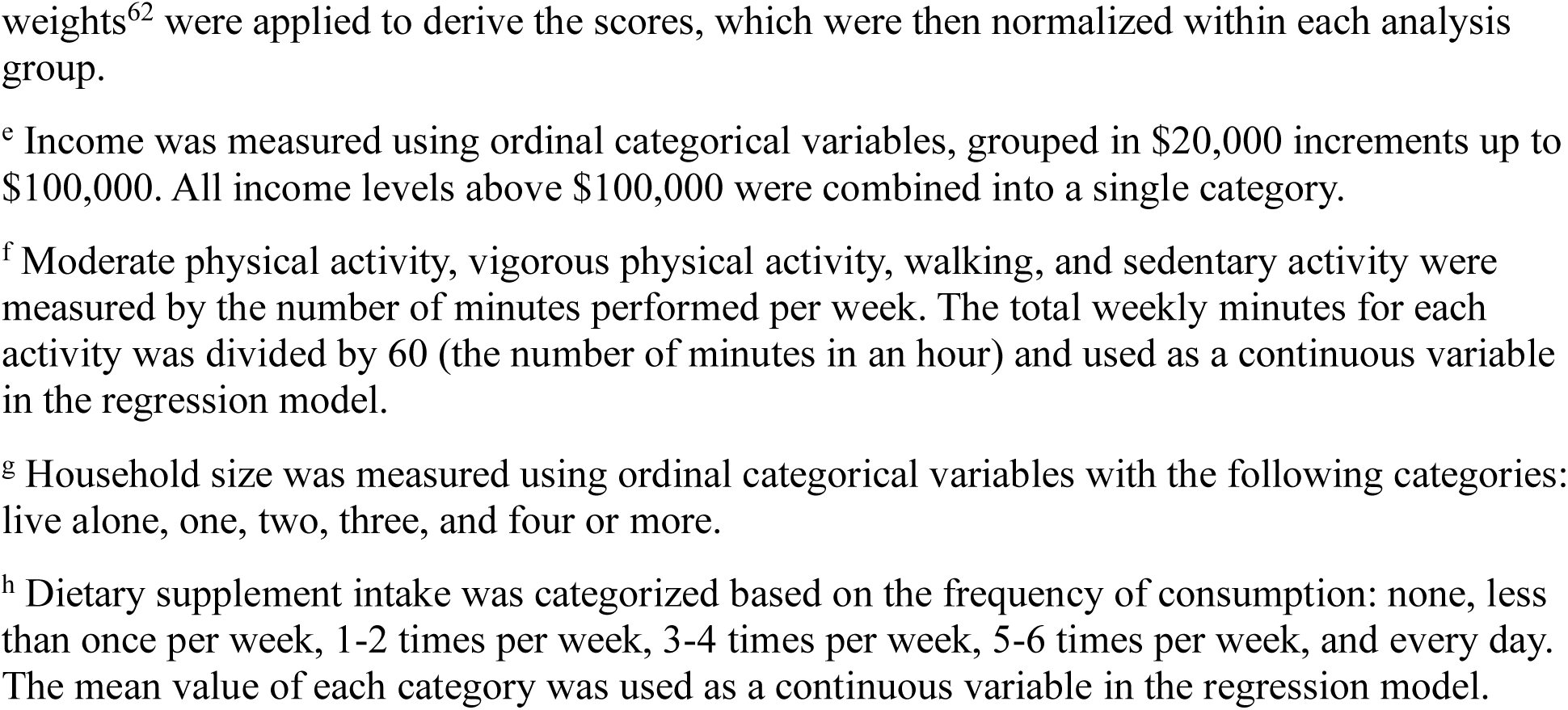
SDOH predictors of RAAS inhibitor-induced angioedema in the GERA cohort^a^.

| Factors | Black Participants<br>(N=617) |  | White Participants<br>(N=15 449) |  |
| --- | --- | --- | --- | --- |
|  | OR (CI) | *P* | OR (CI) | *P* |
| Alcohol Consumption (per Drink per Day)<sup>b</sup> | 1.36 (0.87-2.14) | .17 | 1.28 (1.13-1.46) | <.001 |
| Education<sup>c</sup> | 1.13 (0.81-1.57) | .47 | 0.87 (0.77-0.98) | .023 |
| Western Diet score (per Standard Deviation)<sup>d</sup> | 0.89 (0.58-1.37) | .60 | 0.98 (0.83-1.16) | .83 |
| Prudent Diet score (per Standard Deviation)<sup>d</sup> | 1.19 (0.77-1.82) | .45 | 0.92 (0.77-1.09) | .32 |
| Income<sup>c</sup> | 1.25 (0.84-1.84) | .29 | 1.20 (1.03-1.40) | .022 |
| Marital Status (Effect: Not Married vs. Reference: Married) | 2.80 (1.16-6.77) | .023 | 1.30 (0.88-1.92) | .20 |
| Moderate Physical Activity<sup>f</sup> | 1.06 (0.98-1.14) | .16 | 1.02 (0.99-1.04) | .26 |
| Vigorous Physical Activity<sup>f</sup> | 0.96 (0.92-1.01) | .13 | 1.00 (0.98-1.01) | .74 |
| Walking<sup>f</sup> | 0.98 (0.88-1.08) | .64 | 0.96 (0.92-1.00) | .037 |
| Sedentary Activity<sup>f</sup> | 0.99 (0.95-1.03) | .56 | 1.01 (0.99-1.02) | .52 |
| Size of Household<sup>g</sup> | 1.23 (0.88-1.72) | .23 | 1.08 (0.90-1.30) | .39 |
| Smoking Status (Effect: Ever vs. Reference: Never) | 0.98 (0.44-2.18) | .95 | 1.03 (0.75-1.41) | .85 |
| Multivitamin Supplement<sup>h</sup> | 1.11 (0.97-1.26) | .13 | 1.00 (0.95-1.05) | .93 |
| Vitamin C Supplement<sup>h</sup> | 0.84 (0.66-1.06) | .14 | 0.99 (0.93-1.05) | .74 |
| Calcium Supplement<sup>h</sup> | 0.92 (0.76-1.12) | .40 | 1.01 (0.95-1.07) | .72 |
| Vitamin D Supplement<sup>h</sup> | 0.99 (0.77-1.27) | .92 | 0.97 (0.90-1.05) | .46 |
| Vitamin E Supplement<sup>h</sup> | 1.00 (0.79-1.26) | .99 | 1.05 (0.98-1.12) | .18 |
| Selenium Supplement<sup>h</sup> | 1.28 (0.94-1.76) | .13 | 1.01 (0.91-1.13) | .80 |

CI, 95% confidence interval; GERA, Genetic Epidemiology Research in Adult Health and Aging; OR, odds ratio; RAAS, renin-angiotensin-aldosterone system; SDOH, social determinants of health.
<sup>a</sup> Results were derived from multivariate logistic regression. Covariates included age, sex at birth, and drug class (ACE inhibitors or ARBs).
<sup>b</sup> Alcohol consumption was measured based on the number of alcoholic beverages consumed per week, grouped into categories increasing in increments of 5 (e.g., no drinks, <5 drinks, <10 drinks, ..., 30+ drinks). The mean value within each category was divided by the number of days in a week and used as a continuous variable in the regression model.
<sup>c</sup> Education level was measured using ordinal categorical variables representing the highest level of education attained by participants. The categories included: below high school, high school or GED, technical or trade school, some college, college, and graduate school.
<sup>d</sup> Western (“unhealthy”) and prudent (“healthy”) diet scores were calculated based on survey responses regarding the frequency of consumption of specific food items. Previously published
weights<sup>62</sup> were applied to derive the scores, which were then normalized within each analysis group.
<sup>e</sup> Income was measured using ordinal categorical variables, grouped in \$20,000 increments up to \$100,000. All income levels above \$100,000 were combined into a single category.
<sup>f</sup> Moderate physical activity, vigorous physical activity, walking, and sedentary activity were measured by the number of minutes performed per week. The total weekly minutes for each activity was divided by 60 (the number of minutes in an hour) and used as a continuous variable in the regression model.
<sup>g</sup> Household size was measured using ordinal categorical variables with the following categories: live alone, one, two, three, and four or more.
<sup>h</sup> Dietary supplement intake was categorized based on the frequency of consumption: none, less than once per week, 1-2 times per week, 3-4 times per week, 5-6 times per week, and every day. The mean value of each category was used as a continuous variable in the regression model.

**Extended Data Table 3.**
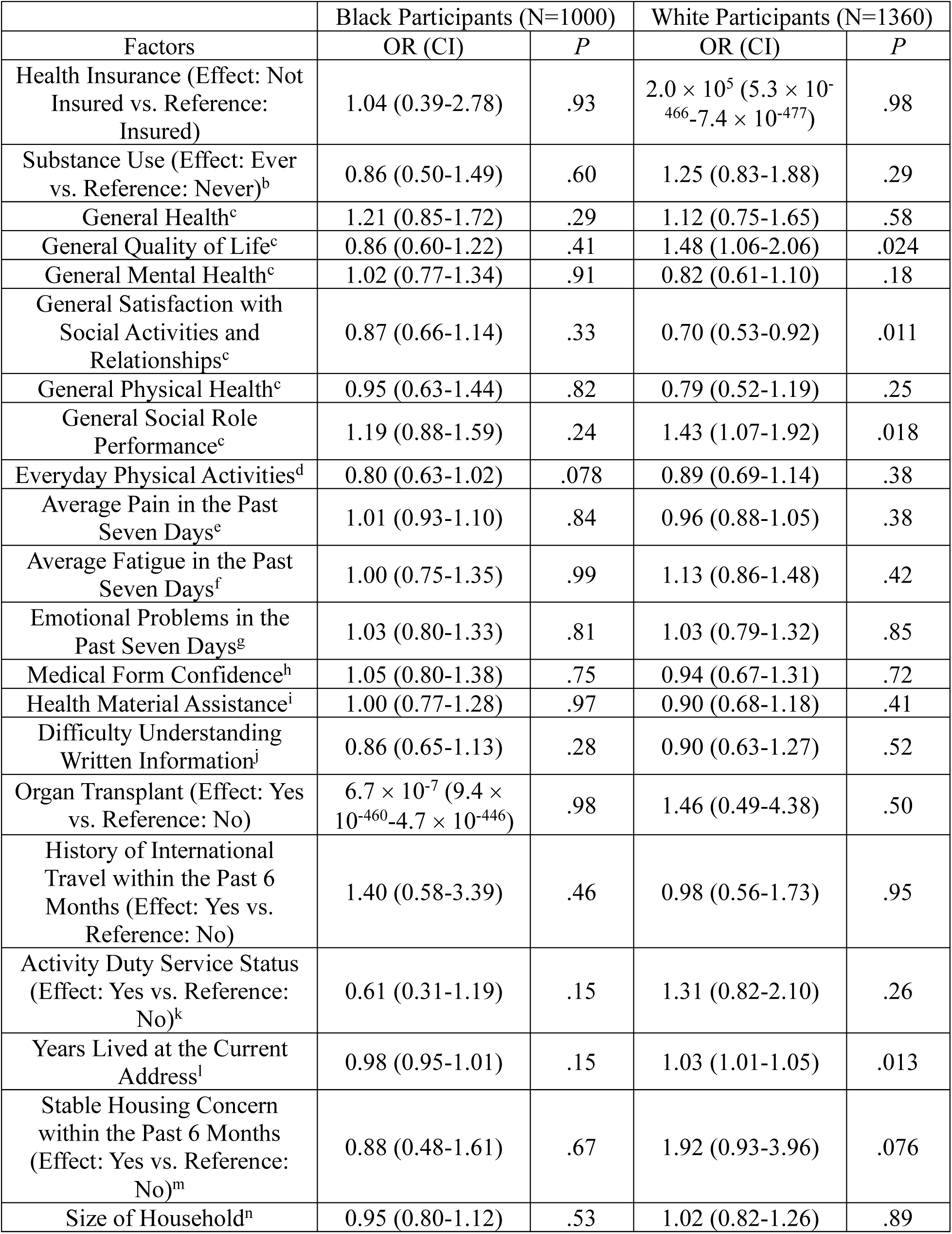
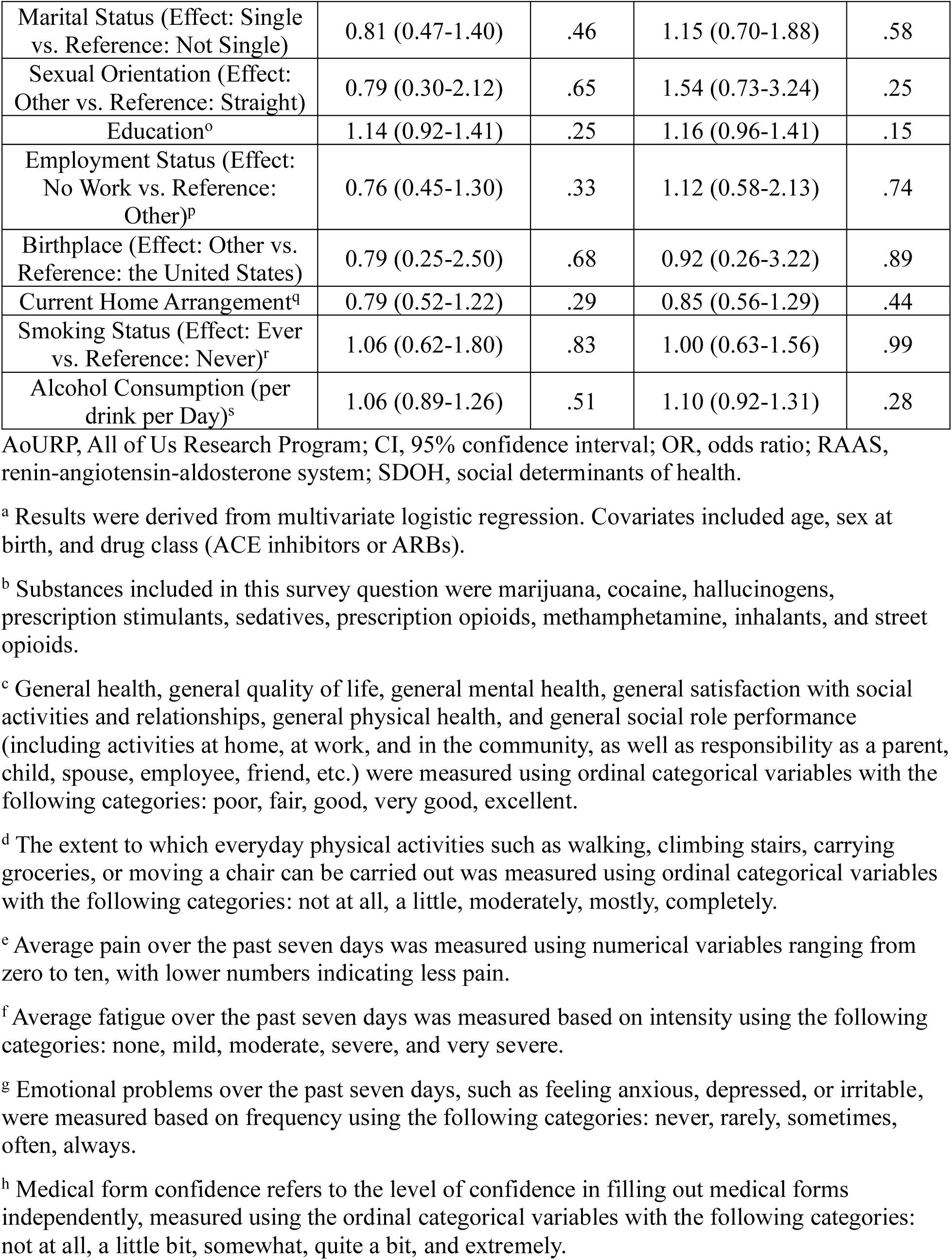
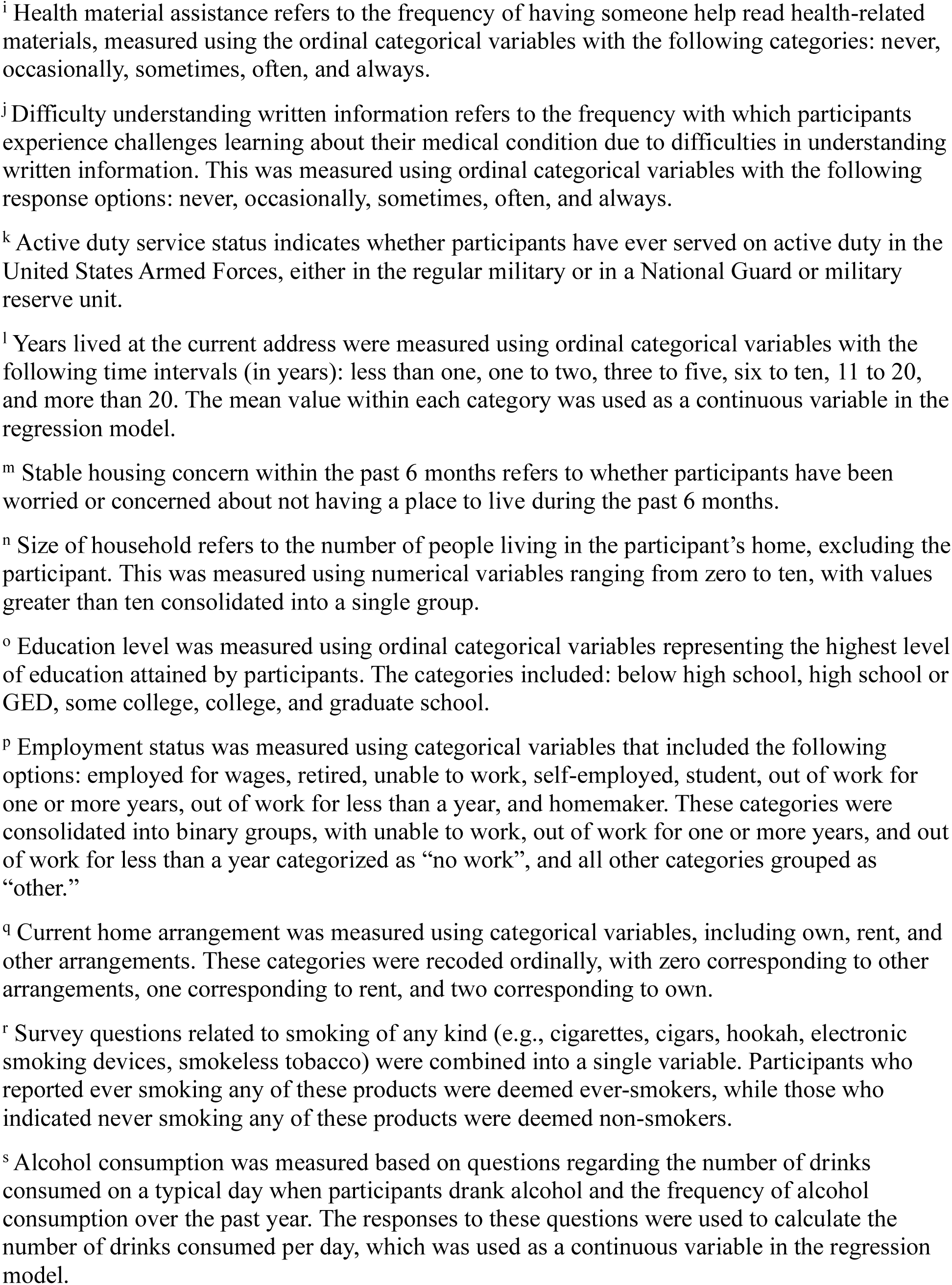
SDOH predictors of RAAS inhibitor-induced angioedema in the AoURP cohort^a^.

|  | Black Participants (N=1000) |  | White Participants (N=1360) |  |
| --- | --- | --- | --- | --- |
| Factors | OR (CI) | P | OR (CI) | P |
| Health Insurance (Effect: Not Insured vs. Reference: Insured) | 1.04 (0.39-2.78) | .93 | $2.0 \times 10^5$ ($5.3 \times 10^{-466}$-$7.4 \times 10^{-477}$) | .98 |
| Substance Use (Effect: Ever vs. Reference: Never)<sup>b</sup> | 0.86 (0.50-1.49) | .60 | 1.25 (0.83-1.88) | .29 |
| General Health<sup>c</sup> | 1.21 (0.85-1.72) | .29 | 1.12 (0.75-1.65) | .58 |
| General Quality of Life<sup>c</sup> | 0.86 (0.60-1.22) | .41 | 1.48 (1.06-2.06) | .024 |
| General Mental Health<sup>c</sup> | 1.02 (0.77-1.34) | .91 | 0.82 (0.61-1.10) | .18 |
| General Satisfaction with Social Activities and Relationships<sup>c</sup> | 0.87 (0.66-1.14) | .33 | 0.70 (0.53-0.92) | .011 |
| General Physical Health<sup>c</sup> | 0.95 (0.63-1.44) | .82 | 0.79 (0.52-1.19) | .25 |
| General Social Role Performance<sup>c</sup> | 1.19 (0.88-1.59) | .24 | 1.43 (1.07-1.92) | .018 |
| Everyday Physical Activities<sup>d</sup> | 0.80 (0.63-1.02) | .078 | 0.89 (0.69-1.14) | .38 |
| Average Pain in the Past Seven Days<sup>e</sup> | 1.01 (0.93-1.10) | .84 | 0.96 (0.88-1.05) | .38 |
| Average Fatigue in the Past Seven Days<sup>f</sup> | 1.00 (0.75-1.35) | .99 | 1.13 (0.86-1.48) | .42 |
| Emotional Problems in the Past Seven Days<sup>g</sup> | 1.03 (0.80-1.33) | .81 | 1.03 (0.79-1.32) | .85 |
| Medical Form Confidence<sup>h</sup> | 1.05 (0.80-1.38) | .75 | 0.94 (0.67-1.31) | .72 |
| Health Material Assistance<sup>i</sup> | 1.00 (0.77-1.28) | .97 | 0.90 (0.68-1.18) | .41 |
| Difficulty Understanding Written Information<sup>j</sup> | 0.86 (0.65-1.13) | .28 | 0.90 (0.63-1.27) | .52 |
| Organ Transplant (Effect: Yes vs. Reference: No) | $6.7 \times 10^{-7}$ ($9.4 \times 10^{-460}$-$4.7 \times 10^{-446}$) | .98 | 1.46 (0.49-4.38) | .50 |
| History of International Travel within the Past 6 Months (Effect: Yes vs. Reference: No) | 1.40 (0.58-3.39) | .46 | 0.98 (0.56-1.73) | .95 |
| Activity Duty Service Status (Effect: Yes vs. Reference: No)<sup>k</sup> | 0.61 (0.31-1.19) | .15 | 1.31 (0.82-2.10) | .26 |
| Years Lived at the Current Address<sup>l</sup> | 0.98 (0.95-1.01) | .15 | 1.03 (1.01-1.05) | .013 |
| Stable Housing Concern within the Past 6 Months (Effect: Yes vs. Reference: No)<sup>m</sup> | 0.88 (0.48-1.61) | .67 | 1.92 (0.93-3.96) | .076 |
| Size of Household<sup>n</sup> | 0.95 (0.80-1.12) | .53 | 1.02 (0.82-1.26) | .89 |
| Marital Status (Effect: Single vs. Reference: Not Single) | 0.81 (0.47-1.40) | .46 | 1.15 (0.70-1.88) | .58 |
| Sexual Orientation (Effect: Other vs. Reference: Straight) | 0.79 (0.30-2.12) | .65 | 1.54 (0.73-3.24) | .25 |
| Education<sup>o</sup> | 1.14 (0.92-1.41) | .25 | 1.16 (0.96-1.41) | .15 |
| Employment Status (Effect: No Work vs. Reference: Other)<sup>p</sup> | 0.76 (0.45-1.30) | .33 | 1.12 (0.58-2.13) | .74 |
| Birthplace (Effect: Other vs. Reference: the United States) | 0.79 (0.25-2.50) | .68 | 0.92 (0.26-3.22) | .89 |
| Current Home Arrangement<sup>q</sup> | 0.79 (0.52-1.22) | .29 | 0.85 (0.56-1.29) | .44 |
| Smoking Status (Effect: Ever vs. Reference: Never)<sup>r</sup> | 1.06 (0.62-1.80) | .83 | 1.00 (0.63-1.56) | .99 |
| Alcohol Consumption (per drink per Day)<sup>s</sup> | 1.06 (0.89-1.26) | .51 | 1.10 (0.92-1.31) | .28 |

AoURP, All of Us Research Program; CI, 95% confidence interval; OR, odds ratio; RAAS, renin-angiotensin-aldosterone system; SDOH, social determinants of health.
<sup>a</sup> Results were derived from multivariate logistic regression. Covariates included age, sex at birth, and drug class (ACE inhibitors or ARBs).
<sup>b</sup> Substances included in this survey question were marijuana, cocaine, hallucinogens, prescription stimulants, sedatives, prescription opioids, methamphetamine, inhalants, and street opioids.
<sup>c</sup> General health, general quality of life, general mental health, general satisfaction with social activities and relationships, general physical health, and general social role performance (including activities at home, at work, and in the community, as well as responsibility as a parent, child, spouse, employee, friend, etc.) were measured using ordinal categorical variables with the following categories: poor, fair, good, very good, excellent.
<sup>d</sup> The extent to which everyday physical activities such as walking, climbing stairs, carrying groceries, or moving a chair can be carried out was measured using ordinal categorical variables with the following categories: not at all, a little, moderately, mostly, completely.
<sup>e</sup> Average pain over the past seven days was measured using numerical variables ranging from zero to ten, with lower numbers indicating less pain.
<sup>f</sup> Average fatigue over the past seven days was measured based on intensity using the following categories: none, mild, moderate, severe, and very severe.
<sup>g</sup> Emotional problems over the past seven days, such as feeling anxious, depressed, or irritable, were measured based on frequency using the following categories: never, rarely, sometimes, often, always.
<sup>h</sup> Medical form confidence refers to the level of confidence in filling out medical forms independently, measured using the ordinal categorical variables with the following categories: not at all, a little bit, somewhat, quite a bit, and extremely.

**Extended Data Table 4.** Significant lead SNVs from a meta-GWAS of RAAS inhibitor-induced angioedema in the Black participant groups^a^.

| Chr | rsID | A1/A2 | FreqA1<sub>Black</sub> | FreqA1<sub>White</sub> | OR (CI) | *P* | Implicated Gene |
| --- | --- | --- | --- | --- | --- | --- | --- |
| 14 | rs72704813 | A/G | 0.20 | 0.22 | 0.64<br>(0.58-0.71) | $4.0 \times 10^{-17}$ | *BDKRB2* |

A1/A2, effect allele/alternative allele; chr, chromosome; FreqA1<sub>Black</sub>, effect allele frequency in the Black participant group; FreqA1<sub>White</sub>, effect allele frequency in the White participant group; GWAS, genome-wide association study; OR, odds ratio; RAAS, renin-angiotensin-aldosterone system; SNV, single nucleotide variant.
<sup>a</sup> Meta-GWAS results reflect the fixed-effects meta-analysis of GWAS summary statistics generated from the All of Us Research Program (AoURP), Genetic Epidemiology Research in Adult Health and Aging (GERA), and Million Veteran Program (MVP) cohorts.

**Extended Data Table 5.** Significant lead SNVs in meta-GWAS of RAAS inhibitor-induced angioedema in the White participant group^a^.

| Chr | rsID | A1/A2 | FreqA1<sub>Black</sub> | FreqA1<sub>White</sub> | OR (CI) | *P* | Implicated Gene |
| --- | --- | --- | --- | --- | --- | --- | --- |
| 14 | rs66515514 | A/C | 0.24 | 0.22 | 0.72 (0.67-0.78) | $4.5 \times 10^{-16}$ | *BDKRB2* |
| 5 | rs26209 | G/A | 0.11 | 0.33 | 1.24 (1.16-1.31) | $1.7 \times 10^{-11}$ | *OTULINL* |
| 15 | rs2903993 | G/A | 0.04<sup>b</sup> | 0.03<sup>c</sup> | 3.99 (2.48-6.43) | $1.2 \times 10^{-8}$ | *CRABP1* |

A1/A2, effect allele/alternative allele; chr, chromosome; FreqA1<sub>Black</sub>, effect allele frequency in the Black participant group; FreqA1<sub>White</sub>, effect allele frequency in the White participant group; GWAS, genome-wide association study; OR, odds ratio; RAAS, renin-angiotensin-aldosterone system; SNV, single nucleotide variant.
<sup>a</sup> Meta-GWAS results reflect the fixed-effects meta-analysis of GWAS summary statistics generated from the All of Us Research Program (AoURP), Genetic Epidemiology Research in Adult Health and Aging (GERA), and Million Veteran Program (MVP) cohorts.
<sup>b</sup> The allele frequency of rs2903993 in the Black participant group reflects data from the GERA cohort.
<sup>c</sup> The allele frequency of rs2903993 in the White participant group reflects data from the GERA and AoURP cohorts.

**Extended Data Table 6.** Significant lead SNVs in meta-GWAS of RAAS inhibitor-induced angioedema in participants with >5% 1KG-AFR-like genetic similarity^a^.

| Chr | rsID | A1/A2 | FreqA1<sub>Black</sub> | FreqA1<sub>White</sub> | OR (CI) | *P* | Implicated Gene |
| --- | --- | --- | --- | --- | --- | --- | --- |
| 14 | rs72704813 | A/G | 0.20 | 0.22 | 0.66 (0.60-0.72) | 2.6 × 10<sup>-17</sup> | *BDKRB2* |

A1/A2, effect allele/alternative allele; chr, chromosome; FreqA1<sub>Black</sub>, effect allele frequency in the Black participant group; FreqA1<sub>White</sub>, effect allele frequency in the White participant group; GWAS, genome-wide association study; OR, odds ratio; RAAS, renin-angiotensin-aldosterone system; SNV, single nucleotide variant.
<sup>a</sup> Meta-GWAS results reflect the fixed-effects meta-analysis of GWAS summary statistics generated from the All of Us Research Program (AoURP), Genetic Epidemiology Research in Adult Health and Aging (GERA), and Million Veteran Program (MVP) cohorts.

## Figures

**Extended Data Figure 1.**
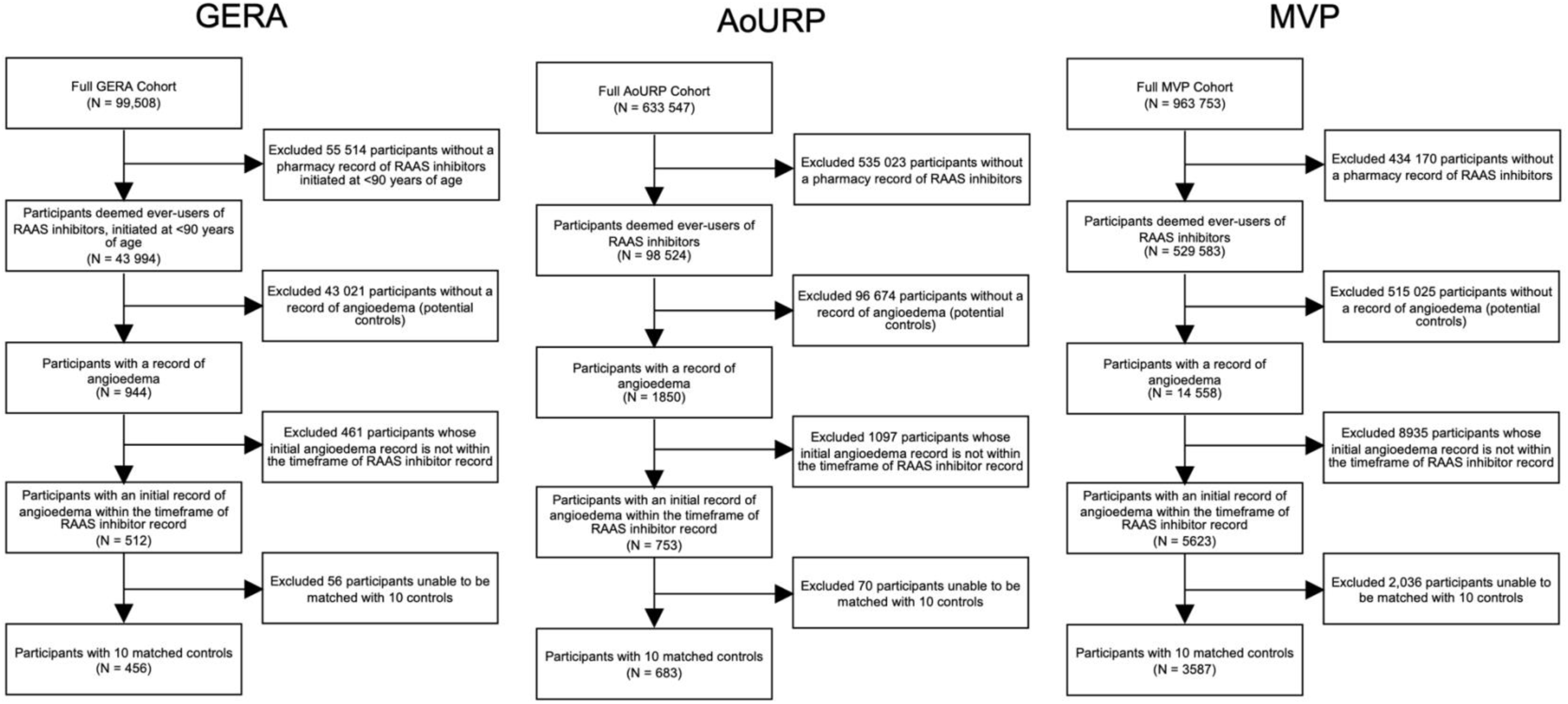
Study inclusion and exclusion criteria for RAAS inhibitor-induced angioedema cases. AoURP, All of Us Research Program; GERA, Genetic Epidemiology Research in Adult Health and Aging; MVP, Million Veteran Program; RAAS, renin-angiotensin-aldosterone system. In the GERA cohort, the timing of RAAS inhibitor initiation was unavailable for participants aged >90 years to protect patient privacy, as the reduced frequency of individuals in this age group increased the risk of identification. As a result, it was not possible to determine whether these participants met the inclusion criteria; therefore, they were excluded from the study. The final matched study population included all eligible cases and their matched controls. In the MVP cohort, some participants were excluded due to withdrawal of their informed consent.

**Extended Data Figure 2.**
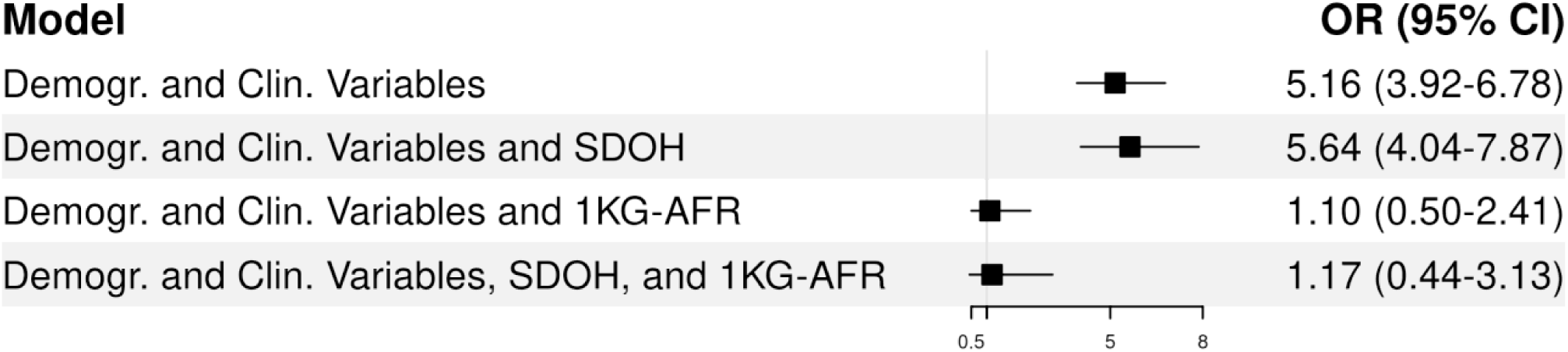
Association of Black race with RAAS inhibitor-induced angioedema risk in the GERA cohort across multivariable logistic regression models 1KG-AFR, 1000 Genomes African superpopulation; clin., clinical; demogr., demographic; GERA, Genetic Epidemiology Research in Adult Health and Aging; RAAS, renin-angiotensin-aldosterone system; SDOH, social determinants of health. The inclusion of SDOH did not account for the observed association with Black race, whereas the addition of 1KG-AFR-like genetic similarity completely abrogated the race effect.

**Extended Data Figure 3.**
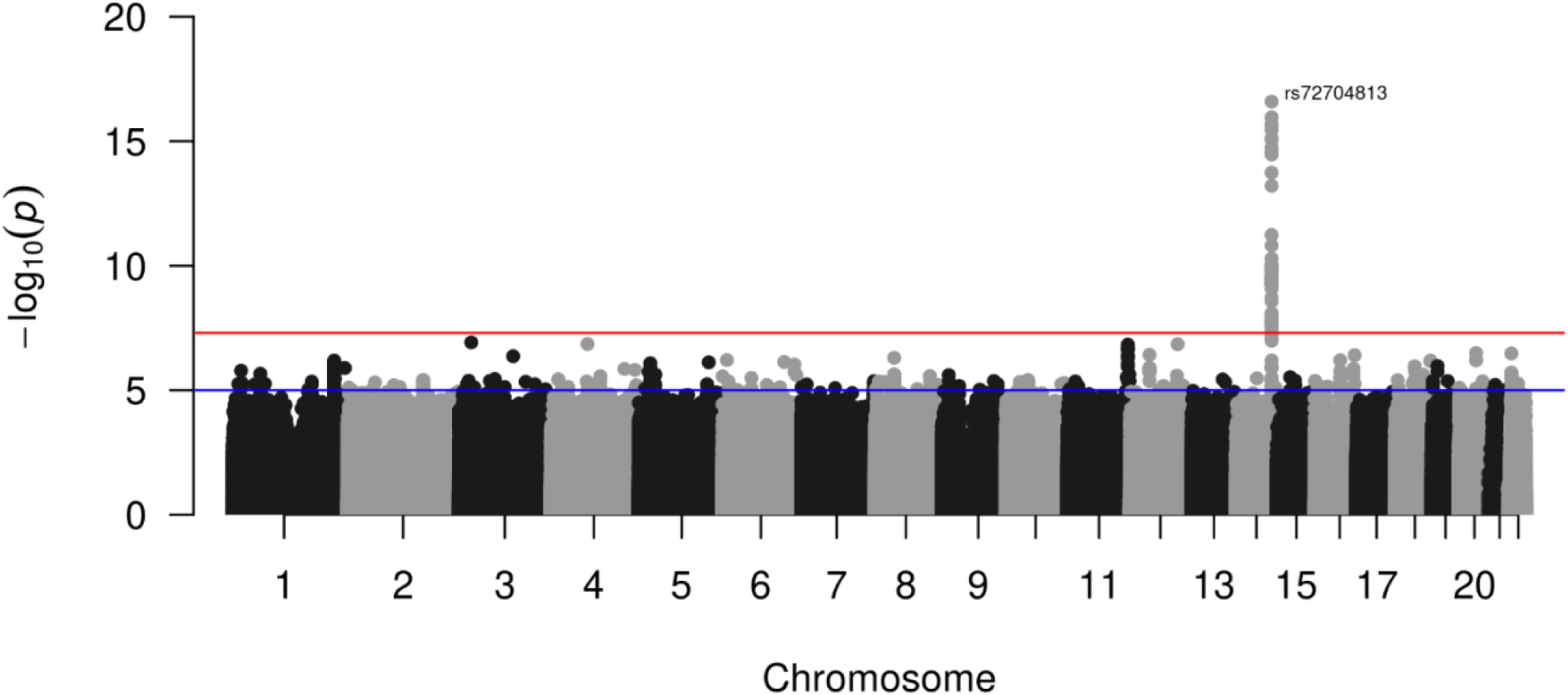
Meta-GWAS for RAAS inhibitor-induced angioedema in participants with >5% 1KG-AFR-like genetic similarity (Case: N = 1790; Control: N = 17 726) 1KG-AFR, 1000 Genomes African superpopulation; GWAS, genome-wide association study; RAAS, renin-angiotensin-aldosterone system. The −log_10_ association *P* values (vertical axis) obtained from a fixed-effects meta-analysis of GWAS summary statistics in participants with >5% 1KG-AFR-like genetic similarity across All of Us Research Program (AoURP), Genetic Epidemiology Research in Adult Health and Aging (GERA), and Million Veteran Program (MVP) cohorts against their genomic positions (horizontal axis) are displayed. The blue horizontal line indicates the prespecified threshold for a suggestive association (*P* = 1 × 10^-5^), and the red horizontal line indicates the threshold for genome-wide significance (*P* = 5 × 10^-8^).

**Extended Data Figure 4.**
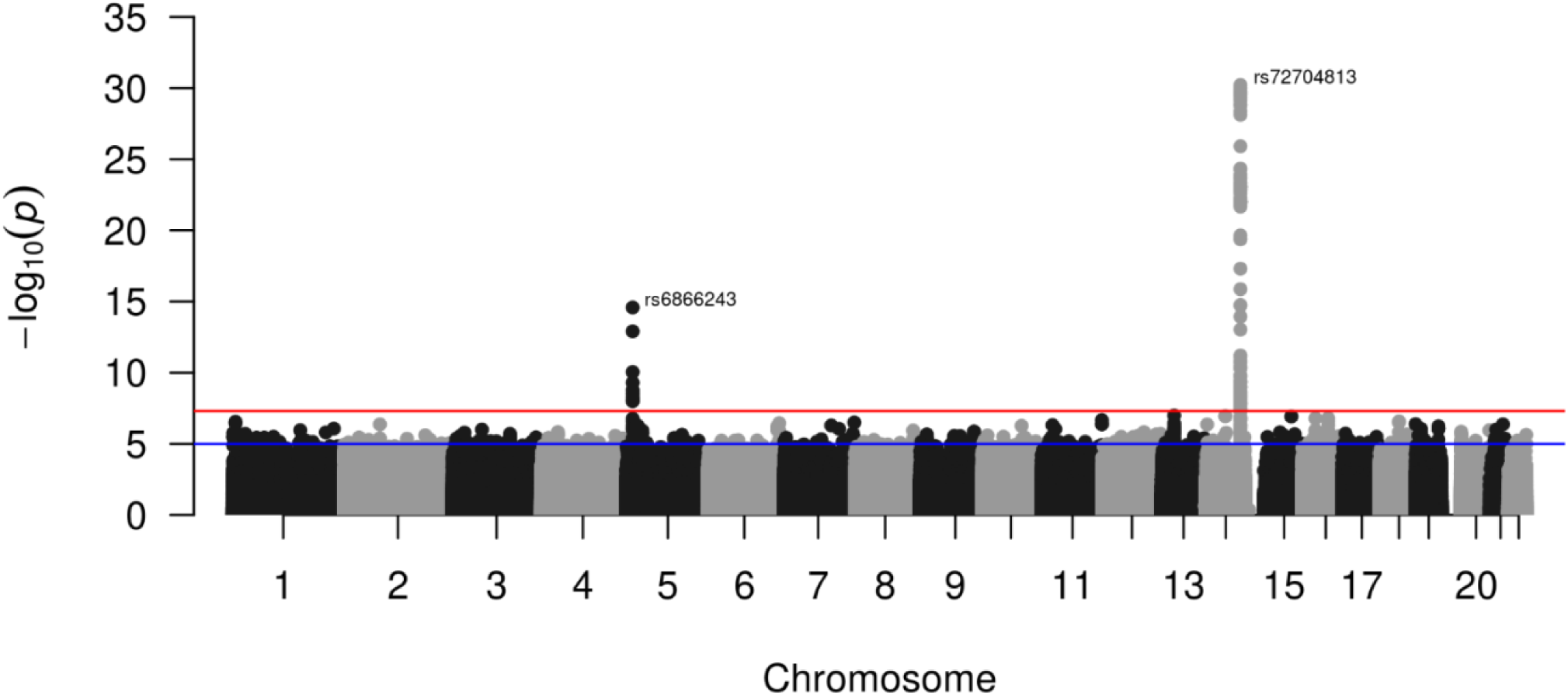
Cross-race meta-GWAS for RAAS inhibitor-induced angioedema. (Case: N = 4461; Control: N = 44 603) GWAS, genome-wide association study; RAAS, renin-angiotensin-aldosterone system. The −log_10_ association *P* values (vertical axis) obtained from a fixed effects, cross-race meta-analysis of GWAS summary statistics across All of Us Research Program (AoURP), Genetic Epidemiology Research in Adult Health and Aging (GERA), and Million Veteran Program (MVP) cohorts against their genomic positions (horizontal axis) are displayed. The blue horizontal line indicates the prespecified threshold for a suggestive association (*P* = 1 × 10^-5^), and the red horizontal line indicates the threshold for genome-wide significance (*P* = 5 × 10^-8^).

