## Supplementary Material for "Genetic and Social Determinants of Renin-Angiotensin-Aldosterone System Inhibitor-Induced Angioedema: A Precision Medicine Health Equity Study"

The Veterans Health Administration (VHA) is a large, integrated health system serving over 9 million enrolled Veterans across the United States^11^. For this study, we accessed EHRs from the Corporate Data Warehouse of the VHA, a repository of clinical data^12^. The EHR data included patient demographics, pharmacy records, and diagnostic codes. The Million Veteran Program (MVP) is a biobank linked to VHA EHRs, comprising approximately 650 thousand participants with genotyping data available^13^. The majority of MVP participants self-identified as White (74%), followed by Black (18%)^14^. Genotyping was performed using a customized Affymetrix Axiom array, followed by quality control. Genomic imputation was conducted using the 1000 Genomes Project (1KG) phase 3 v5 reference panel, with EAGLE v2.4 and Minimac4^15^.

Study Populations and RAAS Inhibitor-induced Angioedema Drug Response

The following classes of renin-angiotensin-aldosterone system (RAAS) inhibitors were eligible for inclusion in our analyses: angiotensin-converting enzyme (ACE) inhibitors, angiotensin-receptor blockers (ARBs), and direct renin inhibitors (DRIs). We included all RAAS inhibitor users; although the strongest effects for angioedema have been observed with ACE inhibitors, there is evidence suggesting that ARBs and DRIs also elevate risk^16^. RAAS inhibitor users, defined as participants who were ever-users of these therapeutic classes, were identified within each cohort using pharmacy records.

PLINK^29^ was used to conduct fixed-effect meta-analyses of the above GWAS summary statistics. The genomic assembly of GWAS results from the GERA cohort was converted from GRCh37 to GRCh38. During this process, variants with identical base pair positions and reference/alternative alleles were removed from the meta-analysis. For variants that shared the same base pair position but exhibited flipped alleles, they were handled as follows: if such variants coexisted in an independent cohort, they were treated as distinct variants. Otherwise, they were treated as a single variant, with adjustments made to account for the allele flip.

To assess whether variants exhibit differential effects between Black and White participant groups for RAAS inhibitor-induced angioedema (i.e., a gene-environment interaction), a genome-wide heterogeneity test was conducted using Cochran’s Q test^30^ on race-conscious summary statistics across all three cohorts. An α = 5 × 10⁻^8^ was used to determine statistical significance.

Polygenic scores were constructed using summary statistics from the heterogeneity test (PS_HET_) and GWAS meta-analysis combining racial groups (PRS_RAAS_) based on MVP data only with PRSice-2^31^. As part of the quality control process, variants with minor allele frequencies (MAF) <0.01, imputation information scores <0.8, duplicate SNVs, and strand-ambiguous SNVs were removed from the base dataset. In the target dataset, variants with MAF <0.01, a genotyping rate <0.1, and a Hardy-Weinberg Equilibrium *P* value (calculated separately for each racial group) <1.0 × 10^-7^ were excluded. Linkage disequilibrium reference data were derived from 1KG-AFR phase 3. Variants for the polygenic scores were selected using *P* value thresholds ranging from 5.0 × 10^-8^ to .5, in increments of 5.0 × 10^-5^. The association between the polygenic scores and RAAS inhibitor-induced angioedema risk (odds) was then assessed by adding these variables to our original logistic regression model of the full AoURP study population (e.g., including clinical and demographic variables).

The above analyses were conducted using R software (R Foundation for Statistical Computing. Version 4.4.2. Available at https://www.R-project.org) or Python (Python Software Foundation. Version, 3.13.2. Available at https://www.python.org) unless otherwise indicated.

25.	National Academies of Sciences, Engineering, and Medicine; Division of Behavioral and Social Sciences and Education; Health and Medicine Division; Committee on Population; Board on Health Sciences Policy; Committee on the Use of Race, Ethnicity, and Ancestry as Population Descriptors in Genomics Research. *Using Population Descriptors in Genetics and Genomics Research: A New Framework for an Evolving Field*. (National Academies Press (US), Washington (DC), 2023).
